# Integrating multi-omics and multi-context QTL data with GWAS reveals the genetic architecture of complex traits and improves the discovery of risk genes

**DOI:** 10.64898/2025.12.19.25342620

**Authors:** Sheng Qian, Kaixuan Luo, Xiaotong Sun, Wesley Crouse, Lifan Liang, Jing Gu, Matthew Stephens, Siming Zhao, Xin He

## Abstract

Recent studies showed that expression QTLs, even from trait-related tissues, explained a small fraction of complex trait heritability. A natural strategy to close this gap is to incorporate molecular QTLs (molQTLs) beyond gene expression, across diverse tissue/cellular contexts. Yet, integrating such QTL data presents analytical challenges. Molecular traits often share QTLs or have QTLs in high LD, complicating the attribution of GWAS signals to specific molecular traits. Our simulations showed that commonly used colocalization and TWAS methods have highly inflated false positive rates in such settings. Building on our earlier work, we developed multi-group causal TWAS (M-cTWAS), for integrating QTLs of different modalities and contexts. M-cTWAS is able to estimate the contribution of each group of molQTLs to the trait heritability, and using such information, identifies the causal molecular traits, informing the modalities and contexts through which genetic variations act on the phenotype. M-cTWAS showed improved control of false discoveries than commonly used methods. Using M-cTWAS, we found that QTLs of multiple modalities greatly increased the explained heritability compared to using eQTLs alone, and enabled the discovery of many more risk genes of a range of complex traits. In conclusion, M-cTWAS effectively integrates diverse molecular QTLs with GWAS to enable causal gene discovery.

## Introduction

Genome-wide association studies (GWAS) have identified a large number of associations for complex traits^1,2^, yet the causal variants, target genes, and underlying molecular mechanisms often remain unknown^3^. A common strategy to interpret GWAS findings is the use of molecular QTLs (molQTLs)^4^. These QTLs associate genetic variants with molecular traits such as expression levels, or RNA splicing. If a GWAS variant of a trait is associated with a molecular trait, say *X*, then *X* likely mediates the effect of the variant on the trait. Building on this idea, many methods have been developed to integrate molQTLs with GWAS^5–7^. This strategy has often used expression QTLs (eQTLs). However, it was estimated that eQTLs from bulk tissues explain relatively small fractions of trait heritability^8,9^. A major challenge in the field is thus to better link the GWAS signals to their molecular effects.

To address this challenge, a natural strategy is to incorporate molecular QTLs (molQTLs) across diverse biological contexts (tissues, cell types and conditions), and modalities. A complex trait often involves interactions of multiple cell and tissue types, thus at the genetic level, the risk variants may act through diverse cellular contexts^10^. Meanwhile, multiple studies have highlighted the importance of expanding modalities beyond eQTLs^11–14^. Splicing QTLs (sQTLs), for example, explained many GWAS signals missed by eQTLs^11^. Epigenetic QTLs, including methylation QTLs (meQTLs) and chromatin accessibility QTLs (caQTLs), explained even larger fractions of GWAS signals than eQTLs from the same tissue and comparable sample sizes^15,16^. QTLs of other molecular traits have been shown to play important roles in complex trait genetics, such as RNA stability, alternative polyadenylation (APA), RNA modification, RNA-editing, and protein levels^12,13,17,18^.

Integrating these heterogeneous QTLs with GWAS, however, presents analytical challenges. Popular methods that integrate eQTLs with GWAS, including Transcriptome-wide Association Studies (TWAS), colocalization (coloc) and Summary-based Mendelian Randomization (SMR), are prone to inflated false positives^6,7,19^. This occurs because eQTLs of a gene, say *X*, may be in linkage disequilibrium (LD) with a causal variant that affects the trait through another mechanism not involving expression of gene *X*. The simulation from our earlier study indicated that TWAS, coloc and SMR have > 50% false positive rates^20^. Integrating over multiple sources of QTL datasets is even more challenging. In such settings, QTLs of multiple molecular traits may be shared or in high LD, complicating the attribution of GWAS signals to the correct molecular trait. Thus naively analyzing each dataset separately and taking the union of results would lead to accumulation of false positives, and incorrect identification of causal molecular traits.

In our previous work, we developed causal-TWAS (cTWAS) to integrate eQTLs with GWAS^20^. cTWAS combines the traditional TWAS with a statistical fine-mapping approach to jointly analyzing all molecular traits and variants in a genomic region. This leads to a much better control of false positive rates than commonly used methods. However, cTWAS was designed to analyze one molecular QTL dataset at a time. Building on cTWAS, we generalized it to multi-cTWAS (M-cTWAS), a scalable framework for integrating multiple QTL datasets across diverse biological contexts and different modalities. M-cTWAS aims to solve two related problems. First, it estimates the enrichment of genetic signals in each group of molQTLs, and their contribution to trait heritability. This is an important problem in practice, as it provides information of the genetic architecture of a trait. This problem is, however, difficult to answer using existing tools, such as coloc or MESC^6,8^. These tools analyze one molQTL dataset at a time, and as a result, can lead to inflated estimates. For example, a researcher may be interested in the contribution of a new type of molQTLs to complex traits. But some of these QTLs may also be eQTLs. Without “controlling” for eQTLs, the study would overestimate the contribution of this type of molQTLs.

Using the estimates of the important genetic parameters, M-cTWAS then addresses the second objective: mapping causal molecular traits of complex traits. From multiple potentially correlated molecular traits of different modalities and contexts as well as genetic variants, M-cTWAS will identify a small number of molecular traits or variants that best explain the genetic associations. This would discover, not only the potential risk gene, but also the molecular modality and context(s) through which the gene affects the trait.

We evaluated M-cTWAS extensively in simulations, and applied it to study the genetics of common traits using multiple molQTLs from GTEx, and from postmortem brains. Our results showed that molQTLs other than eQTLs provide substantial independent contributions to complex trait genetics. M-cTWAS is able to take advantage of diverge molQTLs to discover risk genes of complex traits. Importantly, our analysis highlighted the dangers of applying methods designed for single molQTL dataset such as coloc. Simply taking the union of findings from separate molQTL analysis would lead to high false positive rates of findings.

## Results

### Overview of the M-cTWAS model

We developed multi-group cTWAS (M-cWAS) to analyze molQTLs from multiple contexts and modalities. These molQTLs may be correlated with each other due to LD, or be in LD with other variants in the region. The goal of M-cTWAS is to identify the exact molecular trait(s) that have an effect on a trait of interest, or “causal molecular trait(s)”. To appreciate our problem, imagine a locus has eQTLs of two gene expression traits, denoted as *X*_1_ and *X*_2_, from two tissues A and B, respectively (**Figure 1A**). It also has QTLs of two protein traits *X*_3_ and *X*_4_ from tissues A and B. Among these four molecular traits, only *X*_2_ has a causal effect on the trait *y*. However, multiple correlations complicate the analysis. The eQTLs of *X*_1_ and *X*_2_ are correlated due to LD; and the protein QTLs (pQTLs) of *X*_3_ and *X*_4_ are shared and in high LD with a nearby variant that directly affects *y* (**Figure 1A**). If each molecular trait is tested individually for association with the trait, all four molecular traits would appear to be associated with *y*.

**Figure 1:**
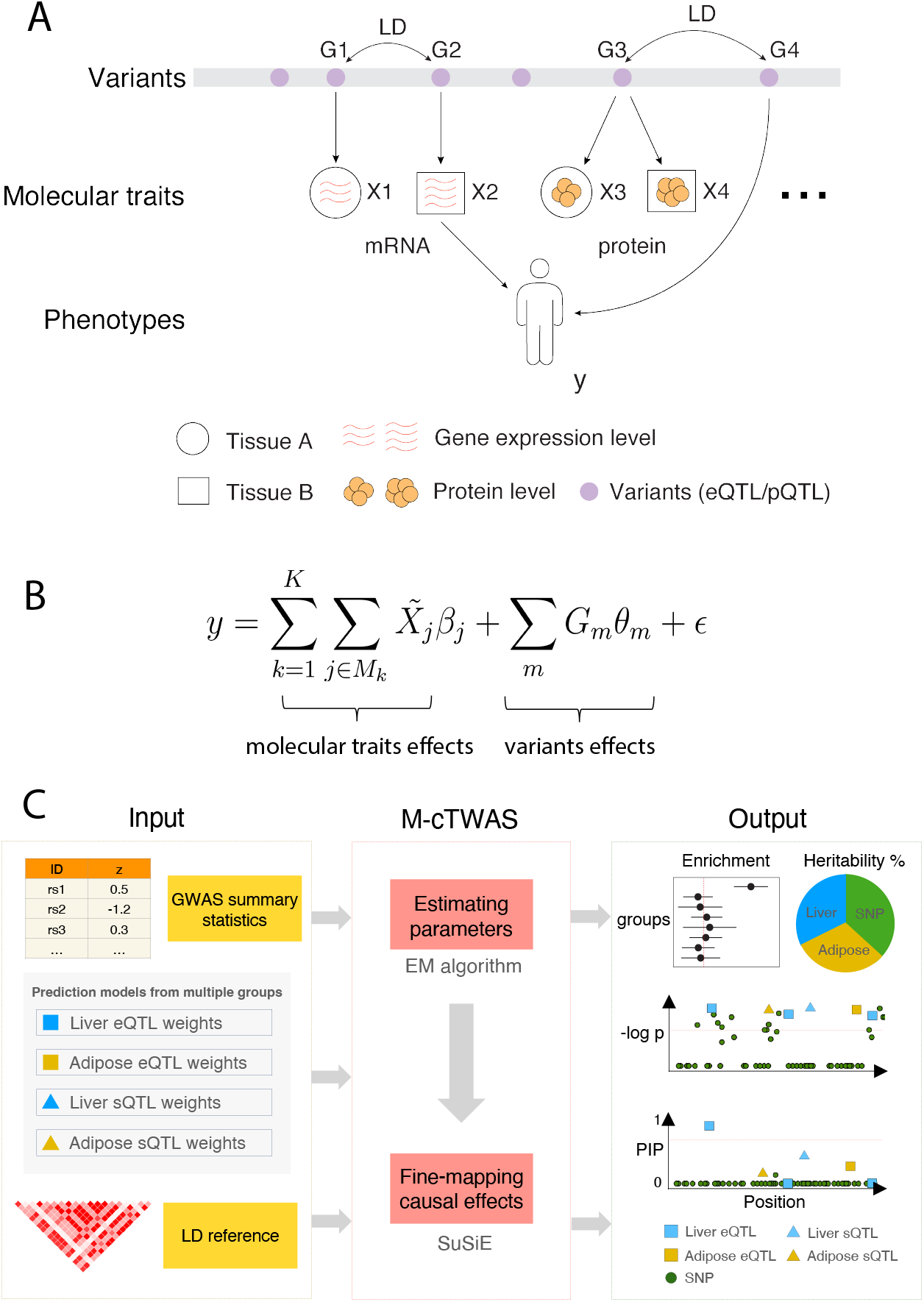
Overview of the M-cTWAS model. **(A)** LD among variants (QTLs) of different molecular traits creates false positives in molQTL analysis. *G*_1_ to *G*_4_ denote genetic variants, *X*_1_ to *X*_4_ molecular traits and *y* the phenotype. Different shapes around molecular traits indicate different tissues. Vertical arrows from “Variants” to “Molecular traits” are molQTL associations. Only *X*2 and *G*4 have causal effects on the phenotype *y*. **(B)** Model of M-cTWAS. 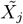 denotes the *j*th predicted molecular trait in group k. *β*_*j*_ denotes its effect size. *G*_*m*_ denotes the genotype of *m*th genetic variant. *θ*_*m*_ denotes its effect size. *y* denotes the phenotype and *ϵ* denotes the residule error. **(C)** The workflow of M-cTWAS. It takes GWAS summary statistics, prediction models of molecular traits and a LD reference panel as input. The inference involves two steps, parameter estimation and fine-mapping of causal effects. The output of M-cTWAS includes estimated parameters and PIPs of molecular traits and variants.

This example suggests that, if we analyze one molecular trait at a time, nearby molecular traits and genetic variants confound the relationship of the tested molecular trait and *y*. M-cTWAS overcomes this confounding problem by jointly modeling the effects of all molecular traits and all variants in a region within a regression model (**Figure 1B**). Similar to TWAS, instead of consider actual molecular traits, we focused here on the genetic component of molecular traits, denoted as 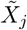 for molecular trait *X*_*j*_. The prediction models of molecular traits from genetic variants are assumed given, in the form of “weights” of variants included in the prediction models. We divided all molecular traits into *K* different groups, representing different modalities or cellular/tissue contexts. It is also important to note that in the regression model, the variant effects refer to their “direct” effects - effects on the trait not mediated through any molecular trait (**Figure S1**).

Solving the regression model, however, is challenging due to the potentially high correlations among variables. To address this problem, we followed the statistical strategy of fine-mapping. We assumed that the causal effects of molecular traits and variants are sparse, and used Bayesian spike-and-slab prior for the effect sizes to introduce sparsity. This “fine-mapping” strategy effectively introduced competitions among correlated variables in the model, to explain the genetic signals on the trait. In fine-mapping, we allowed different prior distributions for variants, and for different groups of molecular traits. Following an Empirical Bayes strategy, these prior distributions are estimated from data. In general, molecular traits are more likely *a priori* to have effects on the phenotype than variants. Similarly, more biologically relevant molecular traits (e.g. those from disease-relevant cell types) would have higher prior than other variables. The use of different prior distributions thus allows us to favor the most relevant molecular traits in the fine-mapping step, hence increasing the power of detecting causal molecular traits.

While our model was formulated with individual level data, we have derived an equivalent model in summary statistics. The input of M-cTWAS then includes summary statistics from GWAS, the weights in the prediction model of the molecular traits, and the reference LD (**Figure 1C**). We estimated the prior parameters from data via the Expectation-Maximization (EM) algorithm using the genome-wide data. Using these parameters, we then performed fine-mapping, one region at a time, on molecular traits and variants using SuSiE^21^ (**Figure 1C**). The output of M-cTWAS is summarized as posterior inclusion probabilities (PIPs) of molecular traits and variants. Under certain assumptions, the PIPs can be understood as the probability of a molecular traits or variants being causal. M-cTWAS also aggregates the PIPs of molecular traits at the gene level. A single gene may have QTLs across different tissue/cell contexts, and may be affected through different molecular mechanisms, e.g. changing expression level or splicing. Thus, combining the evidence of all molecular traits targeting the same gene would increase the power of discovering causal genes. M-cTWAS computes the “gene PIP”, the probability that at least one of the molecular traits targeting a gene has a causal effect (**Methods**).

Although the primary goal of M-cTWAS is to identify causal genes, the estimated parameters in the model’s priors - namely, the proportion of causal effects (*π*) and the effect-size variance (*σ*^2^), provide information on how much genetic effects on the phenotype are enriched in a certain group of molecular traits, and how the heritability is partitioned into contributions from different tissues and molecular modalities (**Figure 1C**). This provides valuable insight into the genetic architecture of complex traits.

M-cTWAS is a generalization of the original cTWAS method. Beyond the main statistical innovations - multiple groups of molecular traits and gene PIP computation, M-cTWAS introduces several other important enhancements. In estimating the genetic parameters, M-cTWAS now provides the standard errors of the estimated values. The software has been optimized to achieve > 10× faster computation and more efficient memory usage. To make it easy to use in the cases where the reference LD may not fully capture the LD in the GWAS samples (in-sample LD), M-cTWAS has included diagnostic tools to detect and correct potential LD-mismatch issues. The full lists of software updates are described in the **Supplementary Notes**.

### M-cTWAS accurately estimates genetic parameters and controls false positive rates in simulations

We designed realistic simulations to evaluate the performance of M-cTWAS. We used genotype data from ∼45,000 samples of White British ancestry in the UK Biobank^22^, restricting to common variants. For molecular traits, we considered gene expression, splicing and RNA stability, with their prediction models (weights) trained from GTEx data^23−25^. Our simulation procedure first imputed molecular traits from genotype data, and then sampled causal SNPs and molecular traits, as well as their effect sizes. We then simulated phenotypes and computed GWAS summary statistics. We considered two main simulation scenarios: (i) the “high correlation” setting using the weights from five GTEx brain tissues - the eQTL effect sizes are highly correlated among these tissues^26^ and (ii) “low correlation” setting using five non-brain tissues, each drawn from distinct tissue groups^8^. In our main results, we focused on the more challenging setting of correlated tissues. In each scenario, two out of the five tissues were designated as causal tissues. In each causal tissue, the proportion of heritability explained (PHE) by gene expression, splicing and RNA stability is set to 5.5%, 2.7% and 0.8%.

We first evaluated the ability of M-cTWAS and other methods to estimate PHE of molQTLs. Both MESC and cTWAS^8,20^ can estimate the PHE from eQTLs, one tissue at a time. However, because eQTLs are often correlated across tissues and with other molecular traits^24,26^, single-tissue eQTL analysis may provide biased estimates. To test this, we applied cTWAS and MESC to estimate PHE of eQTLs in causal and noncausal tissues. In noncausal tissues, which by design contribute zero heritability to the trait, both MESC and cTWAS reported large estimates (**Figure 2A**). These findings underscore the risk of relying on single-tissue eQTL approaches for estimating PHE.

**Figure 2:**
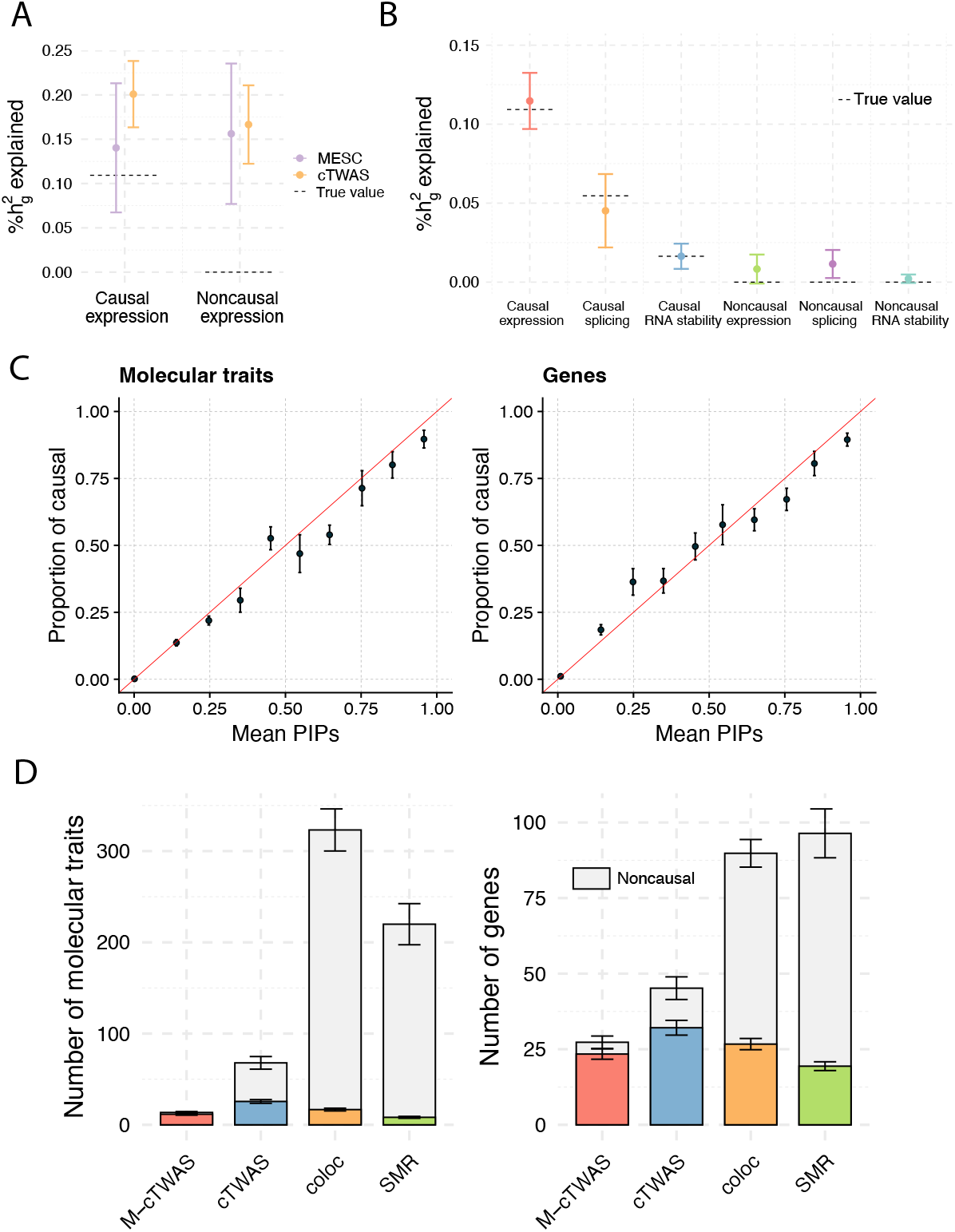
M-cTWAS performance in simulations under the high correlation setting. **(A)** Proportion of heritability explained (PHE) by eQTLs in causal and noncausal tissues estimated by cTWAS and MESC. Dashed horizontal lines indicate the true PHE values. Error bars denote the standard error of the mean. **(B)** Proportion of heritability explained (PHE) across molecular modalities in causal and noncausal tissues estimated by M-cTWAS. Dashed horizontal lines indicate the true PHE values. Error bars denote the standard error of the mean. **(C)** PIP calibration of molecular traits (left) and genes (right). PIPs from all simulations are grouped into bins. X-axis: the average PIPs; Y-axis: the proportion of true causal molecular traits or genes. A well-calibrated method should produce points along the diagonal lines (red). Error bars denote the standard error of the mean. **(D)** Number of molecular traits (left) and genes (right) identified by M-cTWAS, cTWAS, coloc and SMR. We used the following significance thresholds for each method: PIP > 0.8 for M-cTWAS and cTWAS, PP4 > 0.8 for coloc, FDR < 0.05 for SMR with default HEIDI filter. The top gray bars indicate the noncausal molecular traits/genes. The height of a bar is the mean over ten simulations. Error bars denote the standard error of the mean. For the results under the low correlation setting, see **Figures S2**–**S6**.

We then ran M-cTWAS, including all molQTL data in the analysis, to estimate the PHE of each group of molecular traits, and the enrichment of causal effects in each group. The enrichment of a group is defined as the ratio of the proportion of causal molecular traits in this group, and the proportion of causal variants in the variant group. M-cTWAS accurately estimated both PHE and enrichment parameters, with only minor inflation in the non-causal tissues (**Figures 2B, S4**). These results demonstrate that M-cTWAS is able to uncover important genetic parameters.

The main output of M-cTWAS is the PIPs of molecular traits and genes (molecular traits aggregated at the gene level). To properly control the rate of false positive findings, it is important that the PIPs are calibrated, meaning that the average PIP at a certain threshold accurately reflects the true proportion of causal variables. We found that PIPs of both molecular traits and genes are calibrated, especially at the high PIP range (PIP > 0.8) (**Figure 2C**).

Finally, we benchmarked the performance of discovery of causal molecular traits and genes, by M-cTWAS and other methods including cTWAS, coloc, and SMR^6,7,20^. For other methods, we applied each method separately to every molecular trait group (defined by the combination of modality and tissue), and then took the union of the prioritized candidates of all groups. We did not include TWAS, as it is effectively performed within cTWAS and has much higher false positive rate than cTWAS^20^. All these single-group methods exhibited high inflation of false positive rates (**Figure 2D**). In contrast, M-cTWAS effectively controlled false discoveries. Importantly, the power of M-cTWAS, measured as the number of discovered true causal genes, is comparable to other methods (**Figure 2D**).

The results from simulations under the “Low correlation” setting showed similar patterns as reported here (**Figures S2**–**S6**). Altogether, these simulation results highlight the utility of M-cTWAS in estimating important genetic parameters and discovering causal molecular traits and genes.

### M-cTWAS reveals genetic architecture of complex traits

We applied M-cTWAS to study the genetics of 14 commonly studied complex traits, spanning major phenotypic categories such as autoimmune diseases and metabolic traits (**Table S1**). Our molQTL data included gene expression, splicing and RNA stability from GTEx^25^ (**Methods**). We started by dissecting the contributions of molQTLs from different modalities and tissues to the heritability of these traits, and discussed the results of risk gene discovery in later sections.

We first investigate whether the QTLs of different modalities contribute to trait heritability independently. Earlier studies, for example, have suggested that eQTLs and sQTLs have largely independent contributions^11^. For simplicity, we limited this analysis to four traits (LDL, IBD, aFib and SBP) and molQTLs from single tissues, selecting most relevant tissue for each trait (**Supplementary Notes**). We estimated the PHE by gene expression, splicing, and RNA stability, from single modality analysis, i.e. only one type of molQTLs is used each time. These PHE estimates would then be compared with the values from the analyses where all three modalities are included (joint analysis). If all modalities contributed independently, we would expect that PHE of a modality would be similar between separate and joint analysis. We found that the PHE of individual modalities decreased modestly in the joint analysis (**Figure 3A**), suggesting modest sharing among molecular traits within a tissue.

**Figure 3:**
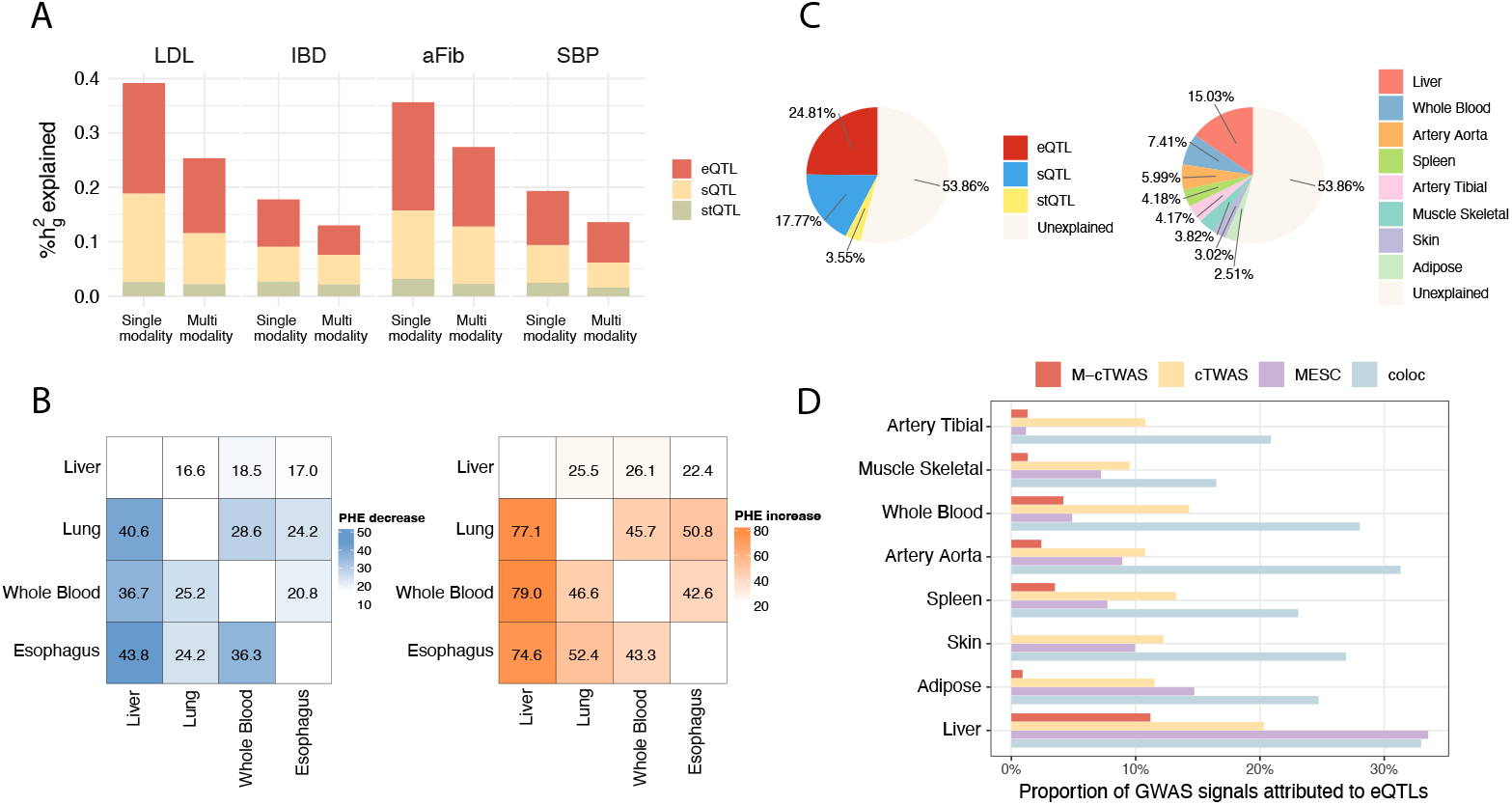
Contribution of molQTLs to trait heritability across molecular modalities and tissues. **(A)** Proportion of heritability explained (PHE) by molQTLs. For each trait, the “Single modality” bar reports PHE of each molecular modality when analyzed separately, and the “Multi modality” bar reports PHE of each molecular modality when analyzed jointly. **(B)** Heatmaps display the percent decrease in PHE attributed to eQTLs in the first tissue after including eQTLs from a second tissue to the model (left), and the percent increase in total PHE attributed to eQTLs in the joint model compared to the PHE of the first tissue alone (right). Each cell corresponds to an ordered tissue pair, where the row tissue is the “first” tissue and the column tissue is the “second” tissue being added. Results are shown for GWAS of LDL. **(C)** Decomposition of trait heritability by molecular modalities (left) and tissue contexts (right) in GWAS of LDL. **(D)** Proportion of heritability explained (PHE) by eQTLs across tissues estimated by M-cTWAS, cTWAS, MESC and coloc. For coloc, we computed the proportion of significant GWAS loci that have colocalized signals with eQTLs (PP4 > 0.8). Results are shown for GWAS of LDL. Notes: in this figure, Esophagus refers to Esophagus Mucosa, Skin refers to Skin Not Sun Exposed Suprapubic and Adipose refers to Adipose Visceral Omentum.

Next, we evaluated cross-tissue sharing by quantifying the change of PHE of eQTL (or sQTL) for a given tissue after incorporating a second tissue. For this analysis, we focused on LDL as the phenotype, and top four tissues ranked by PHE of eQTL in single-tissue analysis. We found that the PHE from gene expression (**Figure 3B**, left panel) or splicing (**Figure S7**, left panel) in the first tissue decreased to varying degrees when a second tissue was included in the model. On the other hand, the joint analysis of two tissues substantially increased the total PHE compared to the first tissue alone (right panels of **Figure 3B** and **Figure S7**). These findings highlight that molecular traits across tissues are partially correlated but not redundant, suggesting the importance of incorporating multiple tissues to capture the full genetics of complex traits.

We then applied M-cTWAS to systematically partition the contributions of three molecular modalities and multiple relevant tissues to trait heritability, including all these groups of molecular traits in a single joint analysis. To simplify the analysis, we limit to selected tissues for each trait (2-8 per trait) (**Table S2, Supplementary Notes**). We found that gene expression, summing over all tissues, explains approximately 5-25% of the trait heritability (**Figure S8**). The proportions of heritability explained by splicing and RNA stability are comparable to expression (**Figure S8**). For example, in LDL, we found that gene expression explains 24.8% heritability, while splicing and RNA stability explain 17.77% and 3.55%, respectively (**Figure 3C**).

In terms of tissue contributions, liver and whole blood emerged as the most important tissues for LDL^27,28^, and whole blood and colon sigmoid were identified as key tissues for IBD^29,30^ (**Figures 3C, S8**).

Lastly, we compared M-cTWAS with other methods that estimate the contribution of molecular QTLs to genetics of phenotypes, including cTWAS, MESC and coloc. For coloc, we calculated the proportion of GWAS significant loci colocalized (PP4 > 0.8) with eQTL signals. Even though this does not estimate the contribution to heritability, it is commonly used in practice to quantify the contribution of molQTLs. We compared the estimated contributions of eQTLs from eight tissues to LDL, one tissue at a time, by all these methods. For M-cTWAS, we included all the tissues and all three modalities in a joint analysis. All methods produced high estimates in liver, consistent with its central role in LDL^27^ (**Figure 3D**). However, single-tissue methods reported high estimates in less relevant tissues, such as skin and muscle, whereas M-cTWAS appropriately shrank these estimates towards zero. Similar patterns were observed for other complex traits (**Figure S9**). In conclusion, to accurately estimate the contribution of molQTLs from some tissues/modalities to a trait, it is important to control for molQTLs from other related tissues and modalities.

### M-cTWAS discovered putative risk genes of complex traits

Having investigated the genetic architecture of complex traits, we evaluated the discovered molecular traits and genes by M-cTWAS in the 14 complex traits. The number of candidate genes identified by M-cTWAS range from 10-250 across traits (**Figures 4A, S10, Table S3**). Compared to cTWAS with gene expression from the top relevant tissue, and M-cTWAS with eQTLs alone, the multi-tissue multi-omics analysis greatly improved the power of discovering candidate genes (**Figure 4A**).

**Figure 4:**
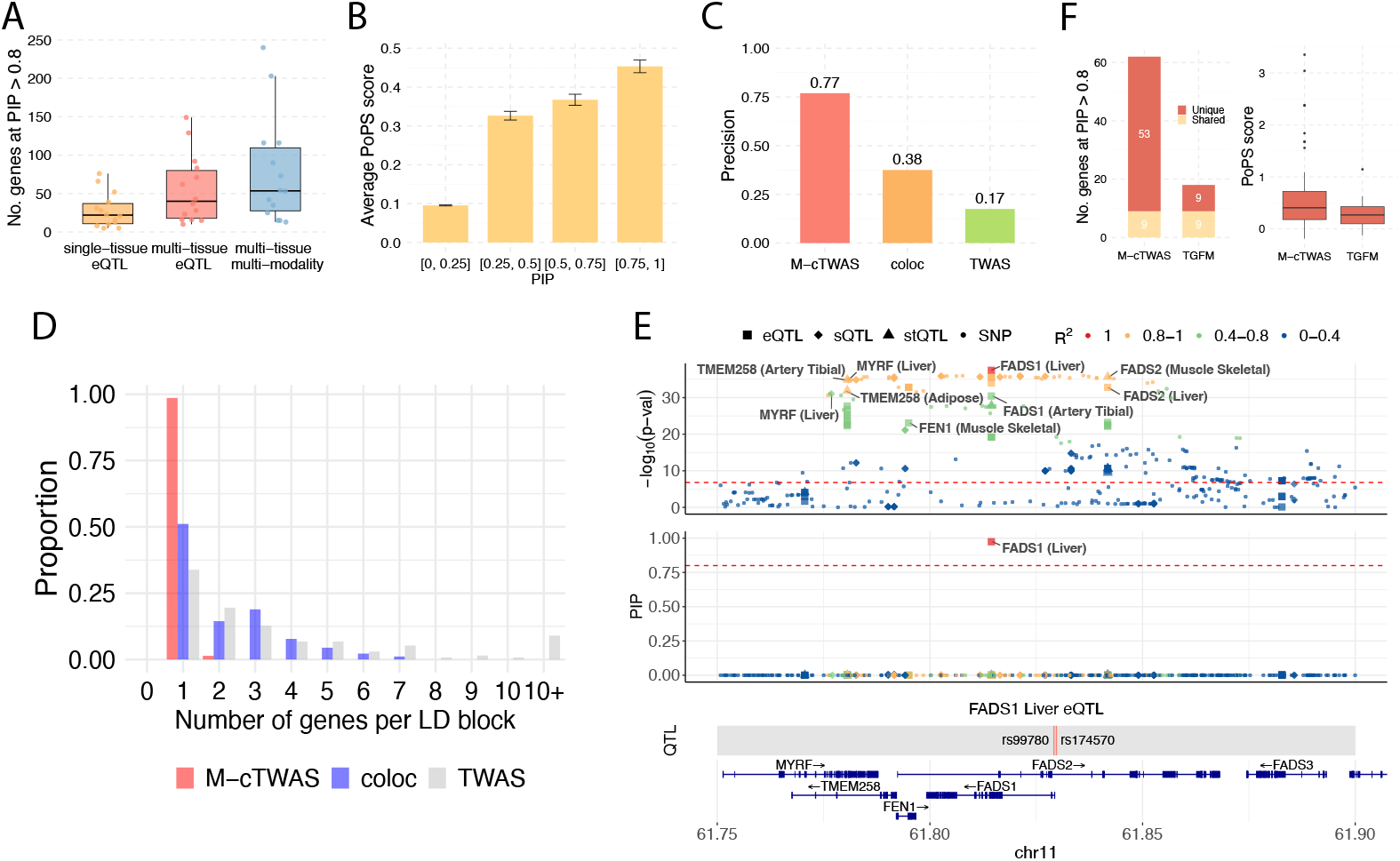
Discovery of risk genes and validation by M-cTWAS. **(A)** Number of candidate genes identified by M-cTWAS across 14 traits in three models: single-tissue eQTL (yellow), multi-tissue eQTL (red) and multi-tissue multi-modality (blue). **(B)** Mean PoPS score (y axis) stratified by M-cTWAS posterior inclusion probability (PIP) bins across 14 traits. Bars show the average PoPS across genes within each PIP bin. Error bars denote the standard error of the mean. **(C)** Precision of M-cTWAS, coloc and TWAS in distinguishing LDL silver standard genes from nearby bystander genes in the same loci. **(D)** The histogram of the number of discovered genes for LDL per LD block, by M-cTWAS, coloc and TWAS. **(E)** Locus plot of the *FADS1* gene locus. The upper panel shows associations of molecular traits and variants with the trait (LDL). The middle panel shows PIPs of molecular traits and variants. The lower two panels show the genomic location of the QTLs (weights in prediction model) of the focal molecular trait and the gene track. Different molecular modalities are shown in different shapes. Colors indicate the LD with the focal molecular trait. Adipose refers to Adipose Visceral Omentum. **(F)** Number of genes identified by M-cTWAS and TGFM in GWAS of LDL at the PIP > 0.8 threshold (left). Colors indicate unique and shared genes identified by two methods. PoPS scores of genes uniquely identified by M-cTWAS and TGFM (right).

To validate the candidate genes identified by M-cTWAS, we first used polygenic priority score (PoPS)^31^. PoPS prioritizes genes by learning trait-relevant gene features including gene expression patterns and associated gene sets such as Gene Ontology terms^31^. Across all traits, genes with higher PIPs tend to have higher PoPS scores (**Figures 4B, S11**).

We next evaluated the results of LDL and IBD more closely, taking advantage of the knowledge of many know risk genes of these traits. We used a list of 69 known risk genes of LDL from literature, denoted as “silver standard” genes^20,32,33^(**Table S4**). In the loci containing silver standard genes, we assessed whether M-cTWAS, and for comparison, coloc and TWAS, can distinguish the silver standard genes from other “bystander” genes in the same loci^20^. We found M-cTWAS achieved 77% precision, i.e. 77% of genes prioritized by M-cTWAS are silver standard genes, close to the theoretical expectation of 80% (based on the PIP threshold). TWAS and coloc, in contrast, have much lower precision (**Figure 4C**). We further assessed performance in IBD using a list of curated high-confidence risk genes (**Table S5**) (**Methods**). M-cTWAS similarly has a high precision of 83%, outperforming coloc and TWAS (**Figure S12**).

As another way to assess the results of M-cTWAS, we inspected the number of genes prioritized by M-cTWAS in individual loci. We reasoned that in most GWAS loci, even when there are multiple causal variants, there is likely a single risk gene^19,20,34^. Thus if multiple risk genes are found in a single locus, it is a sign that some of them are false positive genes. For comparison, we evaluated coloc and TWAS. Given that these methods analyze one molecular QTL dataset at a time, we followed the common practice of taking the union of candidate genes from separate analyses.

We found that M-cTWAS typically prioritized a single candidate gene in most loci (**Figures 4D, S13**). In contrast, coloc and TWAS often implicated two or more genes in a single locus. For example, in LDL, coloc implicated > 1 gene in about half of the loci, and TWAS found even more (**Figure 4D**). As an example, at the FADS1 locus, a number of molecular traits showed strong association with LDL in TWAS (**Figure 4E**, top panel). coloc also implicated five genes supported by 11 SNPs (at PP4 > 0.8), through eQTLs, sQTLs or stQTLs across several tissues (**Figure S14**). Visual examination of two genes confirmed the clear colocalization patterns of the genes with GWAS (**Figure S15**). The colocalization of multiple genes can be explained by the fact that all these 11 SNPs are in strong LD (*R*^2^ > 0.94; **Table S6**), suggesting that it is the LD among QTLs, rather than multiple causal genes, that drives the colocalization of the five genes. M-cTWAS, instead, prioritized a single gene, FADS1 in this locus (**Figure 4E**). FADS1 encodes Fatty Acid Desaturase 1, a key enzyme in fatty acid metabolism and a well-known risk gene of LDL^35,36^. As another example, at the ADAM15 locus in IBD, coloc identified seven candidate genes, driven by a cluster of SNPs in high LD (mean *R*^2^ = 0.77; **Table S7**) (**Figure S16**). Literature search reveals no supporting evidence of these genes. In contrast, M-cTWAS identified a single gene, ADAM15, through splicing (**Figure S17**). ADAM15 is the most biologically plausible IBD gene at this locus. It is up-regulated in IBD and has been implicated in regenerative colonic mucosal differentiation as well as leukocyte transmigration across epithelial and endothelial barriers^37^.

Lastly, we compared M-cTWAS with a recent method, TGFM^38^, which has a somewhat similar goal to ours. TGFM analyzes eQTLs from multiple tissues to identify causal genes and causal tissues. Running TGFM, however, is difficult because it requires individual level genotype data for eQTLs and, in-sample LD for GWAS. It is also very computationally expensive. So we compared M-cTWAS results with published TGFM results. Among the 14 traits we analyzed, we limited to the results of LDL and hypertension (HTN), where the same GWAS data was used by us and by TGFM. For fair comparison, we ran M-cTWAS only with eQTL data. In LDL, M-cTWAS identified several times more genes than TGFM (**Figure 4F**). The results of HTN were similar (**Figure S18**). Moreover, genes uniquely identified by M-cTWAS have higher PoPS scores than those uniquely identified by TGFM (**Figures 4F, S18**). These results thus suggested that M-cTWAS has higher power without losing precision in identifying causal genes than TGFM.

### Novel risk genes provide insights into genetics of complex traits

M-cTWAS not only nominated risk genes, but also provided evidence of specific molecular traits that drive the results. As an approximation, the PIP of a gene is the sum of PIPs of all molecular traits linked to this gene (**Methods**). Thus the results would be informative of the molecular mechanisms (modalities) and the tissues through which genetic variants act on the phenotype. To illustrate this, we summarized the PIP distribution of risk genes of LDL and other traits across modalities and tissues (**Figure 5A, Table S8-9**). We observed that candidate genes are often driven by a single modality and tissue. For example, in the case of LDL, about 1/3 of top genes, such as *LDLR* and *ASGR1*, were driven primarily by sQTLs (**Figure 5A**, left). While stQTLs overall do not make a large contribution, several genes were mostly supported by stQTLs, e.g. *HMGN1* (stQTL PIP = 0.9). A study showed that knockout of HMGN1 in mouse affected the lipid/sterol metabolic pathway^39^. Thus, these results highlighted the importance of considering QTLs beyond gene expression. At the tissue level, while the liver supported the highest number of genes, some genes were primarily supported by other tissues (**Figure 5A**, right).

**Figure 5:**
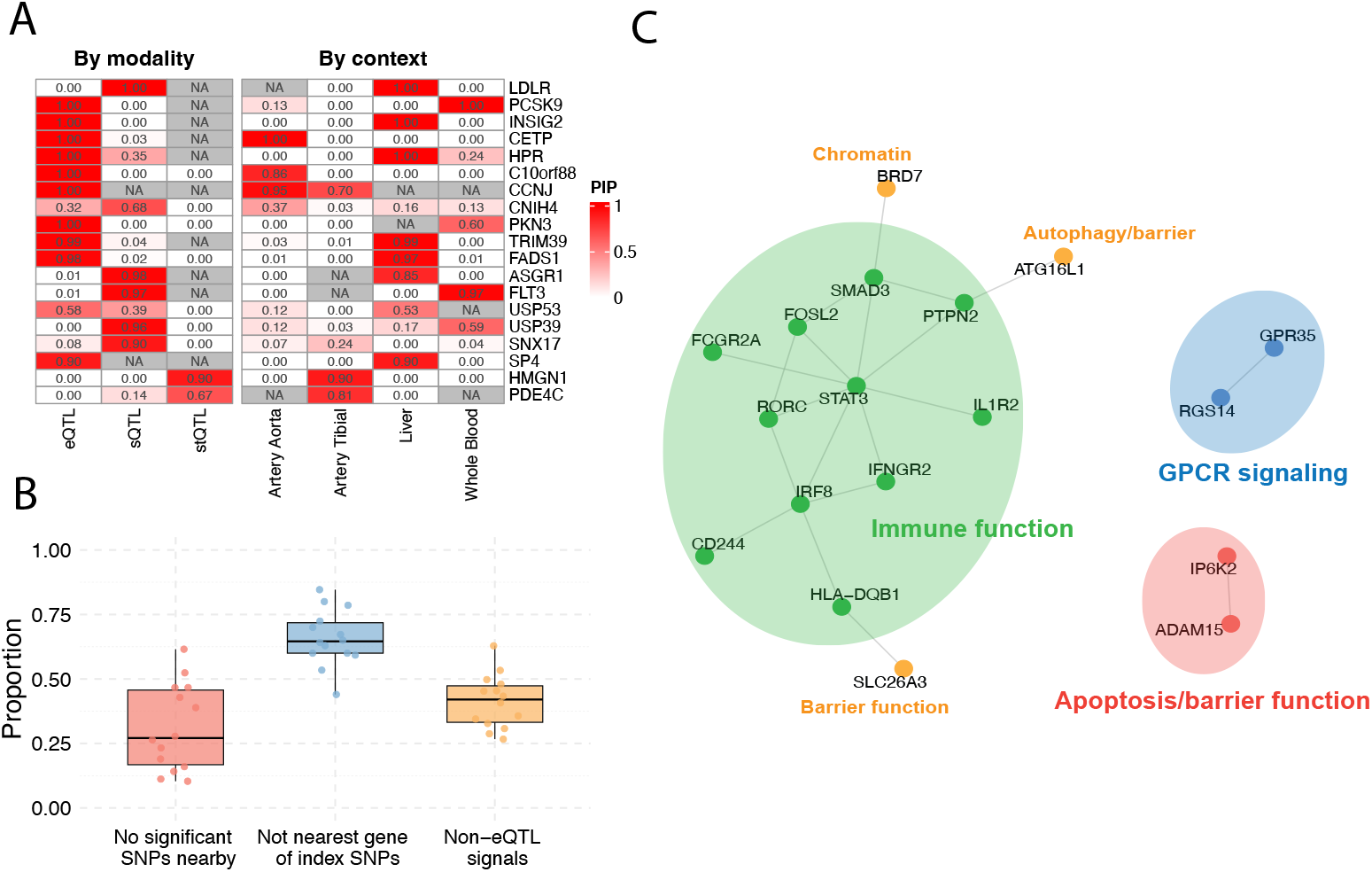
Novel insights into genetics of complex traits. **(A)** PIPs of molecular traits contributing to the top M-cTWAS genes. Left: molecular traits grouped by modalities; Right: grouped by tissue contexts. The top ten genes with highest PIPs and other interesting genes of LDL are included in the heatmap. **(B)** The proportion of genes identified by M-cTWAS across 14 traits that fall into three novel gene categories (i) no significant SNPs nearby: no genome-wide significant SNP (*P* < 5 × 10^−8^) within *±* 500 kb of the gene’s TSS; (ii) not nearest gene of GWAS loci: not the nearest gene to the GWAS lead SNP in a locus; and (iii) non-eQTL signals: genes detected exlusively through splicing and RNA stability. **(C)** Interaction network (STRING) of IBD candidate genes identified by M-cTWAS (PIP > 0.8). Colors indicate cluster memberships from k-means clustering (k=3). Three peripheral genes of the biggest cluster are marked in orange as they are not “core” immune genes.

We believe that the genes discovered by M-cTWAS would allow us to gain novel insights to genetics of complex traits. We first assessed how often the genes found by M-cTWAS were novel, using three criteria: (1) genes located more than 500 kb from genome-wide significant SNPs, (2) genes that are not the nearest gene to the GWAS index SNP, and (3) genes not identified through eQTLs. By these criteria, a large fraction of M-cTWAS genes are novel (**Figures 5B, S19**). Using LDL as an example (**Figure S19**), nearly half (50%) of M-cTWAS genes are not the nearest gene within genome-wide significant loci. Moreover, one quarter (25%) of M-cTWAS genes reside outside significant GWAS loci entirely, while another 25% were discovered through splicing and RNA stability.

We focused on IBD and LDL to illustrate how discoveries of M-cTWAS lead to new understanding of disease genetics. To get a broad picture of the functions of the discovered genes, we performed network analysis using the STRING database^40^. For IBD, we identified a major cluster, associated with leukocyte activation (GO:0045321, FDR = 0.00084), consistent with the immunological basis of the disease^41^ (**Figure 5C**). This cluster contains some known IBD risk genes, including *STAT3, SMAD3* and *ATG16L1* ^42–44^, as well some interesting novel genes. For instance, *RORC* is a master regulator of Th17 cell differentiation, which is central to IBD pathogenesis^45,46^. *RORC* has two major isoforms, of which *RORC2* is preferentially expressed in immune cells^47^. M-cTWAS identified *RORC* through splicing, and the corresponding sQTL associated with the isoform ratio (**Figure S20**). *CD244* modulates inflammatory responses through the MAPK/NF-*κ*B signaling pathway, thus a plausible gene of IBD^48^. Several genes lie in the peripheral of the cluster also look plausible. For example, *SLC26A3* is a crucial colon-specific gene that encodes an intestinal sulfate/chloride transporter, with an important role in maintenance of intestinal epithelial barrier integrity^49^. BRD7 is a chromatin remodeler, and positively regulates type I interferon signaling^50^.

We found two additional groups of genes (**Figure 5C**). One group contains *IP6K2* and *ADAM15*, which are related to apoptotic processes^51,52^. *ADAM15* has been discussed in the previous section and recent work showed that targeting an isoform of *IP6K2* enhanced the intestinal epithelial barrier against inflammation^53^. The second group contains *GPR35* and *RGS14*, genes in the G-protein signaling pathway. Emerging evidence suggests that G-protein signaling plays a significant role in regulating the intestinal mucosal immune system^54^. Several RGS proteins (not including *RGS14*) have been proposed as therapeutic targets in IBD^55^.

For LDL, the genes we found formed into five clusters (**Figure S21**). The largest cluster corresponds to the cholesterol metabolism (GO:0008203, FDR = 1.68 × 10^−8^), and includes well-known LDL genes such as *LDLR, PCSK9*, and *SNX17* ^56−58^. Interestingly, M-cTWAS discovered *LDLR, PCSK9* and *SNX17* through splicing from liver, gene expression from whole blood, and splicing from artery tibial and skin, respectively (**Figure S22**), underscoring the advantage of using molQTLs from multiple tissues and modalites. Another cluster includes *ACVR1C, INHBB* and *DMTN*, which participate in the Activin receptor signaling pathway (GO: 0032924, FDR = 0.0263). We also uncovered a cluster, comprising *USP53, USP39* and *TRIM39*, involved in the ubiquitination processes, pointing to a possible connection between protein turnover and lipid metabolism^59^.

### Brain epigenetic QTLs inform the genetics of Neuropsychiatric diseases

Our analysis so far has been focused on QTLs related to mRNAs. Studies have highlighted contributions of QTLs of epigenetic features (epiQTLs), including chromatin accessibility (caQTLs) and DNA methylation (meQTLs), to genetics of complex traits^15,16^. Nevertheless, epiQTLs are expected to alter gene expression, thus would overlap with eQTLs^60,61^. Dissecting the independent contribution of epiQTLs from eQTL is thus difficult, and has not been rigorously done before. We thus took advantage of M-cTWAS to study the contribution of eQTLs and epiQTLs to neuropsychiatric disease, focusing on Schizophrenia (SCZ)^2^. We jointly analyzed eQTLs from GTEx brain^25^, and meQTLs and caQTLs also from postmortem brains^14,62^.

We found that all three modalities explained comparable proportions of heritability (**Figure 6A**). Altogether, epiQTLs nearly doubled the PHE compared to eQTLs, highlighting the importance of epiQTLs. Notably, the results were similar with a larger brain eQTL dataset from PsychENCODE^63^ (*n* = 1866) instead of GTEx, highlighting the robustness of our findings (**Figure S23**). We next focused on individual cis-regulatory elements (CREs) supported by epiQTLs. We noticed that some epigenetic features are close, e.g. multiple CpGs sites often form clusters, and share epiQTLs^64^. We thus combined such features into clusters, and denoted them as individual CREs (**Methods**). Combining caQTLs and meQTLs, our analysis identified 24 CREs from epiQTLs, compared to 15 genes from eQTLs (**Figure 6B**). Notably, these 24 CREs are not located in the same loci as genes identified by eQTLs, representing novel discoveries that would be missed by using eQTLs alone.

**Figure 6:**
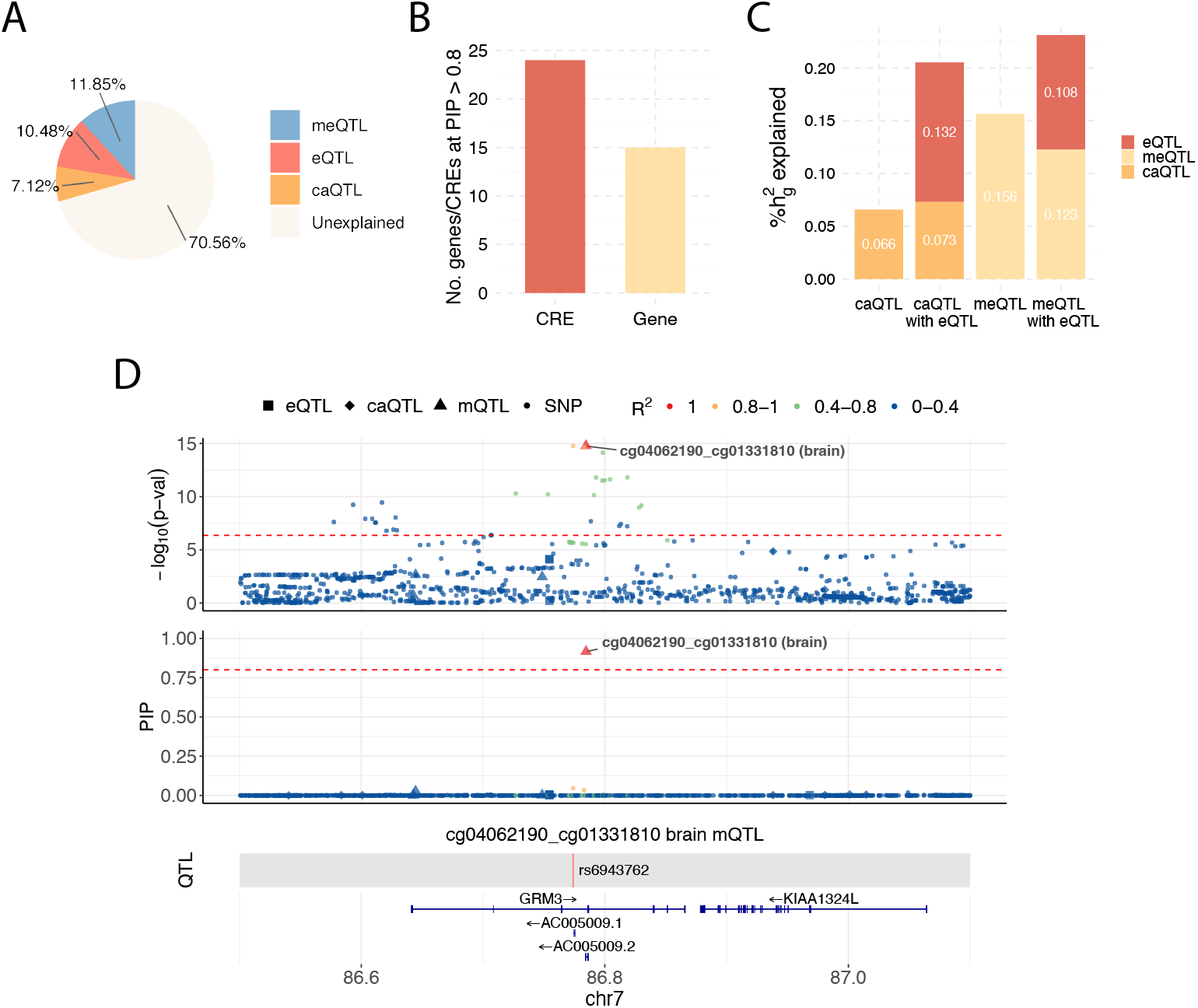
Contribution of brain epigenetic QTLs to schizophrenia. **(A)** Partition of schizophrenia (SCZ) heritability across three types of molQTLs. **(B)** Number of candidate genes and cis-regulatory elements (CREs) identified by M-cTWAS in GWAS of SCZ. **(C)** Proportion of heritability explained (PHE) by epigenetic features (DNA methylation and chromatin accessibility) in the presence and absence of gene expression. **(D)** Locus plot of the *GRM3* gene locus. The upper panel shows associations of molecular traits and variants with the trait (SCZ). The middle panel shows M-cTWAS PIPs of molecular traits and variants. The lower two panels show the genomic location of the QTLs (weights in prediction model) of the focal molecular trait and the gene track. Different molecular modalities are shown in different shapes. Colors indicate the LD with the focal molecular trait. The top molecular trait cg04062190 cg01331810 is a merged CpG site of two nearby CpG sites cg04062190 and cg01331810.

These results were somewhat puzzling given that we expected epiQTLs to primarily function through their effects on gene expression. We thus investigated to what extent epiQTL effects were mediated through eQTLs. We performed M-cTWAS analysis using epigenetic features alone, and compared with the results from the joint analysis where eQTLs were included. Surprisingly, we found that adding eQTLs had little or modest effects on the PHE of caQTLs and meQTLs (**Figure 6C**). At the level of individual CREs, only a small fraction of CREs found using epiQTLs alone had substantially reduced PIPs after including eQTLs (**Figure S24**). These results together suggested that most epigenetic features act independently of GTEx eQTLs.

We investigated two hypotheses that may explain the findings: first, the epigenetic features may have cell-type restricted expression effects, thus missed by eQTLs from bulk tissues; and second, the GTEx eQTLs may be under-powered to detect eQTLs. To assess cell-type specific patterns of CREs, we focused on 242 CpG sites in the 24 CREs found by M-cTWAS. We annotated these CpGs using ATAC-seq data of seven major cell types from PsycheENCODE^65^. 178 CpGs overlapped with Open chromatin regions (OCRs) in at least one cell type. Roughly one-third of these CpGs appeared in all cell types, while another third were present in only one or two cell types (**Figure S25**). This pattern suggests that many CpGs are cell-type specific. To investigate the second hypothesis, we tested whether the top QTL of CpGs and OCRs in candidate CREs were eQTLs in MetaBrain^66^, which has a much larger sample size than GTEx (n = 2,683). Indeed, about 2/3 of meQTLs and caQTLs were confirmed as eQTLs in MetaBrain (**Figure S26**). We note, though, this may be an over-estimate given that some of the epiQTLs may simply be in LD with eQTLs. In summary, we think both hypotheses are plausible and may explain why epiQTLs seem to be largely independent of GTEx eQTLs.

Finally, we highlighted an example of how M-cTWAS analysis with epiQTLs led to novel findings. We identified a CpG site (PIP = 0.92) in the intron of *GRM3* gene. Notably, the gene expression traits near *GRM3* all have low associations with SCZ risk (**Figure 6D**). The CpG site is in the OCR of 4 out of 7 major cell groupings (Astro, Endo, Exc, Oligo) from ATAC-seq^65^, suggesting its regulatory function. Interestingly, the supporting SNP (rs6943762) is not an eQTL in adult brain from multiple sources (GTEx, PEC, MetaBrain), but is an eQTL of *GRM3* in the developing brain^67^. This suggests that epiQTLs from adult brain may capture regulatory effects that are present early in development, but missed in the adult (see Discussion). *GRM3* (metabotropic glutamate receptor 3) regulates synaptic transmission and plays an important role in brain function^68^. While earlier studies have linked genetic variations of *GRM3* with SCZ^69^, there is limited functional support from human genetics. Our results here thus provided evidence supporting this gene and pointed out candidate variant and CRE for future studies.

Altogether, our results highlighted that caQTLs and meQTLs explain genetic signals missed by eQTLs. This may be partially explained by higher sensitivity of epiQTLs in detecting cell-type restricted regulatory effects of genetic variants.

## Discussion

In this study, we developed M-cTWAS, a unified framework for integrating heterogeneous molecular QTL datasets across multiple biological contexts and molecular modalities with GWAS data. M-cTWAS estimates the enrichment and proportion of explained heritability by each group of molQTLs across contexts/modalities. Taking advantage of these parameters, M-cTWAS identifies causal molecular traits through fine-mapping; and causal genes by aggregating evidence of all molecular traits targeting the same genes. Our extensive data analysis highlights the contribution of RNA-related QTLs (sQTL, stQTL) and epiQTLs to the genetics of complex traits. Using these molQTLs helps identify causal genes or regualtory elements missed through eQTL-based analyses alone.

Our findings highlighted the limitations of colocalization analysis. This is probably the most commonly used type of analysis, to estimate the contribution of molQTLs (through percent of GWAS loci colocalized with QTLs) and to discover specific risk genes. We provided several lines of evidence that the results can be substantially inflated. First, our simulations demonstrated that genes discovered by colocalization results have high false positive rates (**Figure 2D**). Secondly, using a set of silver standard genes, we found that coloc results have low precision (**Figure 4C**). Lastly, coloc often identified two or more genes in a single GWAS locus (**Figures 2D, S13**). These results are probably driven by high LD between QTLs of multiple molecular traits, instead of true biological signals.

Several recent methods have also sought to extending the TWAS framework for gene discovery, including GIFT, TGFM, and TGVIS^38,70,71^. GIFT models the genetically predicted expression of all genes in a locus jointly and tests each gene while conditioning other nearby genes. However, it does not model direct variant effects, which are a major source of false-positive TWAS associations^20^. Both TGFM and TGVIS are closely related to cTWAS, in that both use fine-mapping to identify molecular traits, while accounting for direct variant effects. TGFM is designed primarily for multi-tissue eQTL data, without explicitly incorporating other molecular modalities. Additionally, TGFM attempts to account for the uncertainty of prediction weights of gene expression, but the computational strategy relies on repeated sampling of weights and is very expensive. In practice, TGFM seems to be overly conservative, as we have shown (**Figure 4F**). TGVIS jointly models the effects of expression, splicing, and SNPs on a trait, and uses SuSiE to select variables. However, TGVIS does not estimate the parameters of the prior distributions, and treats all molecular traits and SNPs equally. Given that SNPs are usually much more numerous, this may significantly reduce the power of discovering causal molecular traits. Furthermore, without the parameters, TGVIS cannot learn about how heritability is enriched and partitioned among different groups of molecular traits.

Our results highlighted the contribution of epiQTLs, beyond eQTLs, to the heritability of complex traits. Our analysis suggested that epiQTLs may be better powered to detect regulatory effects than eQTLs of comparable samples size. We offered some rationales of this finding. First, expression is generally a more complex trait than epigenetic features. Suppose a variant, say *G*, first affects a CRE, which in turn affects gene expression. Given that the expression of this gene is subject to other influences beyond the CRE, the percent of variance (PVE) of expression explained by *G* would be smaller than PVE of the CRE explained by *G*, leading to lower power of eQTL than epiQTLs. Secondly, epiQTLs in one context may capture eQTL effects in different contexts. This possibility has been suggested by several studies and our own. In GTEx, mQTLs in one tissue are often not eQTLs in the same tissue, but may emerge as eQTLs in other tissue types^72^. In a study of molQTLs in stimulated immune cells, many eQTLs are stimulation-specific, yet they are caQTLs even in the resting state^73^. In our study, the mQTL of GRM3 was discovered in adult brain (**Figure 6D**), yet it was an eQTL only in developing brain, but not adult brain. Altogether, these evidence suggest that epiQTLs may allow us to detect regulatory effects beyond the current cellular or developmental context of study, offering an advantage over eQTL mapping.

We discussed some implications of our findings on future molQTL studies. First, we found that RNA splicing and stability QTLs together made a substantial contribution to heritability, comparable to eQTLs. Indeed, from full-length RNA-seq data, these modalities can be inferred for “free”, so we think it is important to add these QTLs to standard eQTL analysis. Secondly, single-cell RNA-seq is becoming a popular choice for eQTL studies. While this approach improves the detection of cell type/context-specific eQTLs, the commonly used platform captures only 3’ end of mRNAs, limiting the detection of other QTLs, particularly sQTLs. Thus there is a trade-off of using single-cell RNA-seq for mapping eQTLs. Lastly, our results and the discussion above supported a stronger focus on mapping epiQTLs.

We discussed some limitations of M-cTWAS and directions for future work. First, M-cTWAS uses fixed weights in the prediction models of molecular traits, without accounting for uncertainty in the QTL effect size estimation process. Secondly, M-cTWAS is designed for molQTL and GWAS data from the same ancestry. In practice, the ancestries of molQTL and GWAS may be different. Several methods have been proposed recently for multi-ancestry TWAS or fine-mapping^74–77^. We think a similar extension of M-cTWAS framework in the multi-ancestry setting would be an important future direction.

In conclusion, M-cTWAS provides a powerful and scalable framework for integrating multiomics and multi-tissue QTLs with GWAS. As molQTL resources continue to expand, M-cTWAS offers a principled approach to dissecting the genetic architecture of complex diseases and advancing gene discovery.

## Methods

### Multi-group cTWAS (M-cTWAS) model

The M-cTWAS method extends cTWAS framework^20^ to jointly model multiple groups. It incorporates several important features beyond the original cTWAS: computing the standard errors of the model parameters, combining information of multiple molecular traits targeting the same genes, and handling key practical challenges, e.g. LD mismatch between GWAS and reference samples. We start with the main statistical model, and describe these features later and in **Supplementary Notes**.

We assume for now that we have individual level genotype data, phenotypes, and the weights of molecular traits. These molecular traits belong to *K* disjoint groups, e.g. gene expression in several tissues, or different types of molecular traits. From these weights, we can impute the genetic components of molecular traits, denoted as 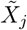 for a molecular trait *X*_*j*_. We thus assume that 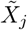‘s are given. We use a linear model to capture how the phenotype, *y*, depends on the genetically predicted molecular traits and genotypes of variants. Formally, let *G*_*m*_ be the genotype of variant *m*, and 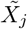 be the *j*-th predicted molecular trait in group *k* (*k* = 1, …, *K*), we have:

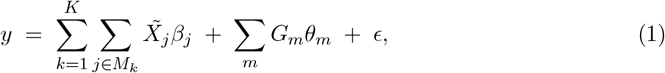

where *M*_*k*_ is the set of molecular traits in group *k, β*_*j*_ and *θ*_*m*_ are effect sizes of the *j*-th predicted molecular trait and *m*th genetic variant, respectively. The error term *ϵ* ∼ *N*(0, *σ*^2^) is assumed to be independent across individuals. All predictors are standardized to unit variance, making effect sizes comparable between groups. Note that the model is effectively a regression model with *K* + 1 groups of variables. For simplicity, in the discussion below, we do not distinguish genetic variants from molecular traits, and consider a general regression model with *K* groups of variables.

We place group-specific spike-and-slab priors on the effect sizes. This form of prior distributions encourages sparsity, thereby addressing the identifiability challenge arising from collinearity among explanatory variables. Also, by specifying different priors for different groups, and learning the prior parameters from data (see below), the model will favor certain groups of variables, e.g. gene expression from an important disease-related cell type, over others in the inference step. This would help identify the variables with true effects.

Specifically, let *γ*_*j*_ be the indicator of whether the *j*-th variable has an effect, our prior for the *j*-th variable from group *k* follows:

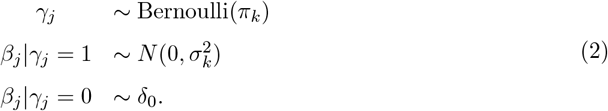

Here, *δ*_0_ denotes the Dirac delta function at zero, *π*_*k*_ represents the prior probability that the *j*th variable from group *k* is causal (i.e., has a non-zero effect on the trait), and 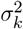 specifies the prior variance of the effect sizes for causal variables within group *k*. While our model here places a different prior for each group, the M-cTWAS software allows several ways to customize the prior. In particular, we allow the variance parameters 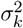’s to be shared among certain groups. For example, one may allow all molecular traits of the same type (e.g. expression from different tissues) to share the same variance parameter.

### Statistical inference of the model

We implemented the M-cTWAS software primarily for summary statistics. The input data consists of (1) the GWAS summary statistics, either *Z*-scores of variants, or estimated effect sizes and their standard errors, (2) the LD among variants, represented by the correlation matrix **R**, (3) the weights of all molecular traits and optionally, the LD information of the variants included in the weights. While this LD information is not strictly required, having the data would make it faster to compute some statistics of the molecular traits (see below).

As the first step, the software computes variant Z-scores (if not provided), and the marginal association (*Z*-scores) of each molecular trait with the phenotype. These associations are effectively statistics from the standard TWAS. M-cTWAS then calculates the correlation of any molecular trait with all other molecular traits and variants, using the LD matrix and the weights. Details of computing *Z*-scores of molecular traits, and the correlation matrix of molecular traits were described in the cTWAS paper^20^.

After computing these necessary statistics, the inference procedure follows two main steps. First, we estimate the prior parameters 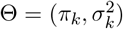 for each group *k* = 1, …,*K* using expectation–maximization (EM) algorithm. Second, using these estimated parameters, we compute the posterior inclusion probability (PIP) for each variable, defined as the posterior probability *P*(*γ*_*j*_ = 1 | *D*, Θ), where *D* stands for the input data.

We maximize the marginal likelihood via an expectation–maximization (EM) algorithm. We first divided the genome into independent LD blocks. Then at each iteration (*t*) of EM, the method performs fine-mapping analysis in each block, given the current parameters *θ*^(*t*)^. For computational reason, we assume at most a single causal effect per block at this step. From fine-mapping, we have the PIP of variable *j*, denoted as 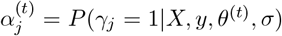; and the the second moment of the posterior effect size of variable *j* given that it has an effect, denoted as 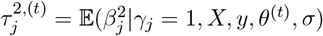. We will then update the parameters:

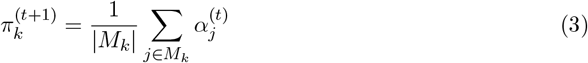

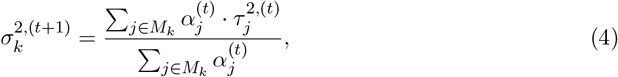

where |*M*_*k*_| is the number of variables in group *k*. We note that EM is done using the entire genome, combining all the LD blocks. Detailed derivation of the EM algorithm is described in the cTWAS paper^20^.

After convergence of EM, we apply SuSiE^21^ within each LD block with estimated prior parameters to compute final PIPs for all variables. At this step, we no longer applied the constraint of at most one causal effect per block.

When performing fine-mapping in both parameter estimation and the final step, we allowed the possibility that a region has no causal signals (i.e. “null model”). In parameter estimation, we used the “single effect approximation” to perform fine-mapping, as described in Equations (17) - (27) in the cTWAS paper^20^. In the final step of fine-mapping causal signals in a region, we set a non-zero null weight in SuSiE to allow the null model. The null weight is chosen so that the prior probability of the null model agrees with that under the single effect approximation, described in Equation (17) in the cTWAS paper^20^.

### Learning about genetic architecture of phenotypes

From the parameters estimated by M-cTWAS, we can compute how heritability is partitioned among different groups of molecular traits and genetic variants. Specifically, the proportion of variance explained (PVE) by the group *k* (including both molecular traits and variants) is given by:

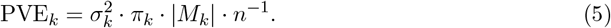

Here |*M*_*k*_| is the group size in group *k* and *n* is the sample size. To obtain the percent of heritability explained (PHE) by the *k*-th group, we divide PVE_*k*_ by the total PVE of all groups.

Additionally, we can estimate how much genetic signals are enriched in molecular traits, compared to variants. We define the enrichment of a group of molecular traits by how often the variables in this group have causal effects, compared to the variant group. Specifically, let |*M*_0_| be the group size of genetic variants, and |*M*_*k*_| the group size of molecular trait group *k*. From M-cTWAS results, we have PIPs of variables in all groups. Note that for this step, we only use PIPs from the parameter estimation step of M-cTWAS, which assumes *L* = 1 (no more than one causal signal per locus) in fine-mapping. We sum the PIPs of all variants, denoted the total as *C*_0_. Similarly, we have the total PIP, *C*_*k*_ of all molecular traits in the group *k*. The total PIP of a group can be viewed as an approximation of the number of causal effects in that group. We then have the log enrichment as:

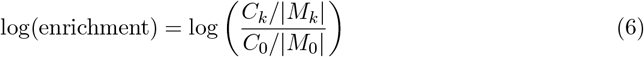

Its standard error can be obtained based on the standard 2 x 2 table analysis (group size and total PIP of the two groups):

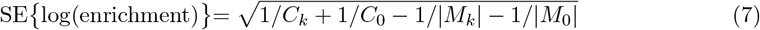

We used Fisher’s Exact Test to obtain the statistical significance of enrichment. We note that *C*_0_ and *C*_*k*_ are not necessarily integers, so we round these numbers in the test.

### Computing gene level PIPs

Multiple molecular traits may target the same gene. Integrating evidence across all such traits can enhance the power to identify candidate genes. We compute the gene PIP, denoted as PIP_*j*_, the probability that at least one of the molecular trait of gene *j* has a causal effect.

In the first step, we aggregate the posterior inclusion probabilities (PIPs) of all molecular traits linked to the gene within each credible set. Let PIP_*jm*_ be the PIP of the molecular trait *m* linked to gene *j*, the total evidence of gene *j* from the *k*-th credible set is given by:

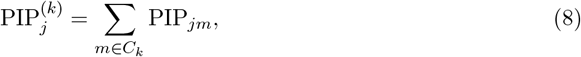

where *C*_*k*_ means all variables in the *k*-th credible set.

In the second step, we aggregate the evidence of gene *j* across all credible sets:

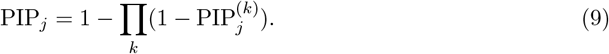

When there is a single credible set, the PIP of a gene is the sum of PIPs of all molecular traits linked to this gene. When there are multiple credible sets, the gene-level PIP would be smaller than the total PIP of all molecular traits linked to gene *j*. As an approximation, if 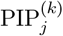’s are small, the PIP of gene is approximately the sum of 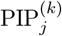’s (expanding Equation 9 and ignoring the higher order terms).

### Prediction models of molecular traits in GTEx

We used gene expression and splicing models from the PredictDB database (https://predictdb.org) based on GTEx v8 data (mashr weights). For RNA stability, we constructed the prediction models from GTEx samples, following the procedure described in **Supplementary Notes**.

### Simulation procedure

We used genotype data from the UK Biobank^22^, limited to individuals of self-reported White British ancestry. After applying standard quality control filters, including removal of samples with missing information, sex mismatches, outliers flagged by UK Biobank, and related individuals, the dataset consisted of *n* = 45, 087 individuals and 6,228,664 SNPs with minor allele frequency > 0.05 and genotyping rate ≥ 95%. The molecular traits and their prediction models were taken from GTEx, as described above. We used two sets of tissues in simulations, based on the extent of correlation of eQTL effects across tissues. (1) Highly correlated tissues include brain cerebellum, brain cortex, brain nucleus accumbens, brain caudate and brain cerebellar hemisphere. (2) Lowly correlated tissues include whole blood, liver, heart left ventricle, stomach and lung.

SNP genotypes were harmonized between prediction models and the UK Biobank data to ensure consistent reference and alternate alleles. We further removed strand-ambiguous SNPs. After the filtering steps, we down-sampled to 5,000,000 SNPs (keep SNPs in prediction models). We imputed gene expression, splicing and RNA stability for all samples using the prediction models. Among the five tissues included in the simulation, we selected two tissues as causal. We then sampled causal molecular traits and SNPs from two causal tissues according to prior inclusion probabilities *π*_*k*_, and their effect sizes were drawn from the corresponding prior variance parameters 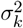. Phenotypes *y* were simulated under the M-cTWAS model defined in Equation (1). The prior parameters *π*_*k*_ and 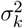 were selected to reflect realistic genetic architectures. Specifically, we set the phenotypic variance explained (PVE) by gene expression, splicing, and RNA stability in each causal tissue to be 2%, 1%, and 0.3%, respectively. And SNP PVE is set to 30%. For simplicity, we fixed the effect size variance of causal molecular traits and SNPs to be 0.02.

To run M-cTWAS, we performed GWAS of the simulated phenotype to obtain summary statistics of SNPs 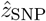. We then ran M-cTWAS with a reference LD calculated with 10% of UKBB samples. For benchmarking, we compared M-cTWAS against cTWAS, coloc and SMR by analyzing 15 molecular trait datasets (spanning five tissues and three molecular trait types). For each of the other methods, we took the union set of candidate genes identified across all molecular traits for comparison.

### GWAS summary statistics

We obtained publicly available GWAS summary statistics from various resources. Specifically, summary statistics for LDL cholesterol (LDL), systolic blood pressure (SBP), inflammatory bowel disease (IBD), atrial fibrillation (aFib), white blood cell count (WBC), schizophrenia (SCZ), were accessed via the IEU OpenGWAS project^78^. GWAS summary statistics for body mass index (BMI), height, platelet count (PLT) red blood cell count (RBC), hemoglobin levels (HB), hypertension (HTN), and type 2 diabetes (T2D) were downloaded from the Pan-UK Biobank project^79^. GWAS data for type 1 diabetes (T1D) were from the GWAS Catalog^80^.

### Running M-cTWAS using GTEx data

We applied the following M-cTWAS settings for all analyses using GTEx weights (Figures 3-5). We used the same weights for gene expression, splicing and RNA-stability as described in the section “Prediction models of molecular traits in GTEx” in Methods. For LD reference, we also used the same LD reference from UK Biobank, as described in the Simulation section. We first estimated the model parameters. We assumed a common prior variance parameter across all groups (including SNPs). We ran up to 50 EM iterations to estimate prior parameters. Regions potentially harboring causal signals were then selected using two criteria: (1) at least one SNP with p-value < 5 × 10^−8^, and (2) total PIP for molecular traits (non-SNP PIP) exceeds 0.5. These regions were then used in the fine-mapping step. We computed correlation among all variables (SNPs and molecular traits) and fine-mapped with SuSiE (L = 5) causal effects in each LD block with estimated parameters. We computed gene PIPs from these fine-mapping results. We performed several post-processing steps, including LD mismatch diagnosis (see below) and resolving the cases where a gene spans more than one block. If the variants of molecular traits of a gene cross more than one block, the PIP of the gene may be unreliable, and this may also affect other nearby genes. So we identified such genes with PIP > 0.5, merge the regions containing molecular traits of these genes, and the re-run fine-mapping (L = 5) on the merged regions.

### Dealing with LD mismatch

Using an out-of-sample LD panel may create LD mismatch in some regions and variants, leading to potential problems in fine-mapping^81,82^. To diagnose potential LD mismatches, we focused on the regions that have some candidate molecular traits of interest. We found these regions with the total PIP from all molecular traits > 0.2. We followed the LD-mismatch diagnostic approach outlines in SuSiE-RSS^81^. For each variant in these regions, we inferred the expected statistic of the variant, based on the Z-scores of nearby variants and the LD of the focal variant and nearby variants. The expected statistic then is compared to the observed statistic of that variant. A significant discrepancy between the expected and observed values indicated a potential LD mismatch. We applied a mismatch threshold of 5 × 10 ^−6^ to flag problematic variants. We then identified the molecular traits affected by these problematic variants and re-performed fine-mapping with SuSiE using the option, *L* = 1, without the LD matrix. When there is at most one causal signal in a region, the fine-mapping results are independent of LD^83^.

### Estimation of the contribution of molQTLs to trait heritability

We run MESC with pre-computed expression scores from GTEx v8 (https://github.com/douglasyao/mesc/wiki/Download-expression-scores) to estimate trait heritability mediated through gene expression in each tissue. For M-cTWAS and cTWAS, we estimated the heritability attributed to molecular traits using the estimated model parameters - see the section “Learning about genetic architecture of phenotypes”.

### Benchmarking M-cTWAS against other methods

We evaluated the performance of M-cTWAS through both simulation studies and real data analyses, benchmarking it against several widely used approaches, including TWAS, coloc, SMR, and cTWAS. These comparison methods are single-group approaches that analyze one molecular dataset at a time. We applied each method separately to every molecular dataset used by M-cTWAS and then combined their results by taking the union of significant findings. For TWAS, we computed Z-scores of molecular traits following the S-PrediXcan^84^ formula internally by M-cTWAS and converted to *p*-values. Statistical significance was determined using a Bonferroni correction adjusted for the total number of molecular traits across all datasets. For coloc, we used full summary statistics from each molecular QTL dataset and considered molecular traits with PP4 > 0.8 as colocalized. For SMR, we applied the method to summary statistics from each molecular QTL dataset (only significant SNPs are included). We considered molecular traits with FDR < 0.05 from SMR with default HEIDI filter as significant findings. For cTWAS, we analyzed each molQTL group separately and considered molecular traits with PIP > 0.8 as causal.

### Validation of M-cTWAS genes

To validate genes prioritized by M-cTWAS, we used two complementary approaches: (1) comparison with PoPS scores^31^, a popular gene prioritization method, and (2) evaluation against known high-confidence gene sets. For the PoPS-based validation, we obtained the complete set of features provided by the PoPS study (https://github.com/FinucaneLab/pops) and used UK Biobank (UKBB) genotype files (bfiles) as input to run PoPS for each trait. For genes with both M-cTWAS posterior inclusion probabilities (PIPs) and PoPS scores, we grouped genes into predefined PIP bins and calculated the average PoPS score within each bin. This analysis was performed separately for each trait and then aggregated across all traits to evaluate the relationship between M-cTWAS gene-level probabilities and PoPS-derived biological relevance.

For the high-confidence gene sets, we used a curated set of “silver standard” genes. For LDL, we used 69 known LDL-associated genes^20,32,33^. For IBD, we used DeepSeek V3.1 to identify high-confidence IBD genes. Specifically, we ran model three times with the prompt “Give me 20 high-confidence IBD risk genes based on literature. Limit to protein-coding genes” and selected genes that appeared in all three runs. To reduce variability, we set the temperature parameter to 0, which yielded 14 IBD genes with strong literature support. For each silver-standard or AI-curated gene, we defined a set of “bystander” genes located within 1 Mb of the corresponding gene as negative controls. We then assessed the ability of M-cTWAS to distinguish silver-standard genes from bystander genes and compared its performance against TWAS and coloc. Precision is defined as the number of detected silver standard genes/(number of detected silver standard genes + number of detected bystandard genes). Detected silver standard genes and bystandard genes are defined at PIP > 0.8 threshold.

### M-cTWAS analysis with epigenetic QTLs

Summary statistics for DNA methylation QTLs in the brain were obtained from Qi et al.^62^, and chromatin accessibility QTLs in neurons and glia were obtained from Zeng et al.^14^. Because access to individual-level genotype data is often restricted, we built simplified prediction models by selecting the most significant SNP associated with each CpG site or ATAC-peak. In these single-SNP models, the weight assigned to the SNP is arbitrary and does not influence the results. Running M-cTWAS under this framework is conceptually similar to a colocalization analysis but with additional adjustment for nearby genetic confounders (see Supplementary Notes of the cTWAS paper^20^). To reduce redundancy, CpG sites located within 1 kb of each other were merged, and the most significant SNP among the merged CpGs was retained in the prediction model. We notice that sometimes two or more CpG sites or ATAC peaks may share the QTLs. In such cases, M-cTWAS will not be able to resolve the causal one, and the evidence is in fact split among these sites/peaks. This would reduce the number of high PIP findings. To address this issue, we merge all CpG sites and ATAC peaks within the same credible set, and denote them as a single cis-regulatory element (CRE). The PIP of the CRE is the sum of PIPs of all sites/peaks.

### Other procedures

For the analysis of pairwise PHE, the percent decrease in PHE is calculated as (PHE_tissue1_ − PHE_tissue1,joint_)*/*PHE_tissue1_ × 100% and the percent increase in total PHE is calculated as (PHE_joint_ − PHE_tissue1_)*/*PHE_tissue1_ × 100%. For the analysis of the number of genes per locus, we only ran coloc in regions with significant TWAS *p*-values. For the analysis of TGFM comparisons, we obtained TGFM results from https://doi.org/10.7910/DVN/S26PFI. For the analysis of novel genes, we defined genes not identified through eQTLs as those detected by the multi-tissue and multi-omics M-cTWAS analysis but not by the multi-tissue eQTL M-cTWAS analysis. For the string network analysis, we used the web server (https://string-db.org) to construct interaction networks.

## Data Availability

All data produced in the present study are available upon reasonable request to the authors.

## Data and code availability

Genotype data from UK Biobank are available through the UK Biobank data access process (http://www.ukbiobank.ac.uk/register-apply/). Publicly available prediction models for 49 GTEx tissues from PredictDB (https://predictdb.org/post/2021/07/21/gtex-v8-models-on-eqtl-and-sqtl/). GWAS summary statistics are available from the IEU OpenGWAS project (https://gwas.mrcieu.ac.uk/), the Pan-UK Biobank project (https://pan.ukbb.broadinstitute.org/downloads/index.html) and the GWAS Catalog (https://www.ebi.ac.uk/gwas/home). Our software is available at https://xinhe-lab.github.io/multigroup_ctwas/.

## Acknowledgments

This work was supported by the National Institutes of Health (NIH) under grants R01MH110531, R01HL163523, R01AI175554 and U19AI162310 (to X.H.). We thank H. Im (University of Chicago), X. Liu (University of Chicago), F. Luca (University of Chicago) and other members of the He Lab for helpful comments on the work and the manuscript.

## Contributions

X.H. conceived the idea and supervised the project. S.Q. and X.S. performed the data analyses. K.L., S.Q. and W.C. improved and implemented the software. S.Q., K.L., X.S. and X.H. wrote the manuscript. S.Z., X.H. and M.S. developed the original cTWAS method and algorithm. M.S. provided advice on technique issues of the method. S.Z. provided feedback on the manuscript. L.L. and J.G. tested the software and provided feedback.

## Supplementary Notes

### Major updates in M-cTWAS software

We made several major improvements in M-cTWAS software. First, we extended the cTWAS framework to incorporate prediction models from multiple molecular trait types across diverse biological contexts. Second, we redesigned the software interface to improve modularity, implementing key M-cTWAS tasks as standalone components. Users may run the full M-cTWAS analysis via the main function or execute individual steps independently. In addition, we implemented a “cTWAS-light” (“no-LD”) mode, that enables M-cTWAS analyses without requiring an LD reference panel. Other important updates include:

### Input processing

- Supports multiple prediction model formats, including PredictDB, FUSION, and top- QTL.
- Provides streamlined preprocessing tools to harmonize GWAS summary statistics, prediction models, and LD reference.
- Offers utility functions for constructing LD matrices from individual-level genotype data and for loading LD matrices of various formats, with flexible support for user-defined custom formats.

### Computation

- Introduces new parameter estimation procedures that enable sharing of prior effect variance parameters across multiple groups of molecular traits. Several schemes of parameter sharing are supported, including: shared variance for all groups of variables (including SNPs); shared variance for all molecular traits; shared variance for all molecular traits of the same type/modality.
- Compute standard errors and *p*-values of enrichment parameters
- Implements new methods to screen genomic regions with likely causal molecular trait signals. This speeds up the computation, as only such regions will be subject to full fine-mapping (allowing more than one causal signal).
- Re-implements a simplified single effect regression (SER) model from SuSiE, incorporating the new implementation of null configuration. It is used in EM to speed up computation.
- Improves computational efficiency by more than ten-fold, enabling large-scale analyses across many molecular traits. A typical M-cTWAS analysis using molQTLs from multiple groups could be completed within 1–3 hours.

### Creating the output

- Introduces new methods to combine evidence from all molecular traits targeting the same genes.
- Enhances visualization of M-cTWAS results, including updated locus plots with integrated gene tracks and fine-mapping result panels.
- Incorporates LD-mismatch diagnosis from SuSiE-RSS in post-processing.
- Introduces a region-merging step to resolve the “cross-region” challenge, where the genetic variants contributing to a molecular trait spans two regions.

### Creating prediction models for RNA stability

RNA phenotypes (RNA stability and alternative polyadenylation usage) and corresponding covariate data were obtained from Munro et al.^1^ (available at https://pantry.pejlab.org/). Genotype data were acquired from GTEx v8^2^, with missing genotypes imputed using Beagle^3^. To control for confounding factors, we performed covariate regression on both RNA phenotypes and genotype matrices, followed by scaling to ensure data comparability across samples.

For each tissue type, we restricted our analysis to RNA molecules previously included in Munro et al.’s prediction models. Samples were randomly partitioned into training (80%) and testing (20%) subsets to facilitate model validation. For each RNA molecular trait, we selected genetic variants located within *±*50 kb of the transcription start site (TSS). We implemented the SuSiE fine-mapping approach (with L = 5) to identify causal variants. From each credible set, we selected variants with the highest posterior inclusion probability (PIP) when multiple variants were present in a credible set. Effect sizes for selected variants were estimated using multiple linear regression, with the resulting coefficients representing SNP effect sizes for prediction models.

We evaluated predictive performance using the coefficient of determination (*R*^2^), calculated as: 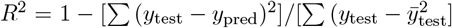, where *y*_test_ represents observed RNA levels, *y*_pred_ denotes predicted values, and 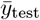 is the mean observed value in the test set. Predictions were aggregated across all cross-validation folds to generate global *R*^2^ values for each RNA molecular trait.

Following cross-validation, we trained the final prediction model using all available samples to maximize statistical power for effect size estimation. Since both RNA stability and alternative polyadenylation contribute to overall RNA regulation, we implemented a complementary modeling approach. RNA stability measurements were used as the baseline predictive feature, while alternative polyadenylation usage data were incorporated as complementary features when they mapped to genes not already represented in the RNA stability dataset. The adjusted and scaled RNA phenotype matrix and genotype matrix were used for QTL mapping. For each molecular trait, we applied the <monospace>lm()</monospace> function in R to perform linear regression and compute *p*-values for all cis-SNPs located within *±*50 kb of the TSS.

### Tissue selection for M-cTWAS analysis in real data

Complex traits typically involve risk genes that exert effects across multiple tissues. To reduce computational burden and minimize human error in tissue selection, we developed an automated procedure to identify relevant tissues for M-cTWAS. Specifically, we utilized splicing sQTL data to select tissues contributing significantly to trait heritability. Compared to eQTLs, sQTLs are more tissue-specific, enabling more precise identification of tissues that play biologically meaningful roles in the genetic architecture of each trait. This approach ensures that M-cTWAS focuses on the most functionally relevant tissues for each phenotype.

To implement this procedure, we first excluded GTEx tissues with fewer than 200 samples to ensure adequate statistical power. We then removed tissues deemed irrelevant to the traits under investigation, including testis, nerve tibial, ovary, prostate, uterus, vagina, and breast mammary tissue. After these filtering steps, 33 GTEx tissues remained, comprising 8 brain tissues and 25 non-brain tissues. We jointly ran M-cTWAS using sQTL weights from all tissues. For type 2 diabetes (T2D) and body mass index (BMI), analyses included all 33 tissues. For other non-psychiatric traits, analyses were restricted to the 25 non-brain tissues, whereas for psychiatric traits, only the 8 brain tissues were included. We ranked tissues based on *p*-values from enrichment tests and retained those passing a Bonferroni-corrected significance threshold (0.05 divided by the total number of tissues tested). The statistically significant tissues were subsequently used in the M-cTWAS analyses. For single-tissue analysis, we used the most relevant tissue of each trait defined as the tissue in which eQTLs explains the highest heritability.

## Supplemental Figures

**Figure S1:**
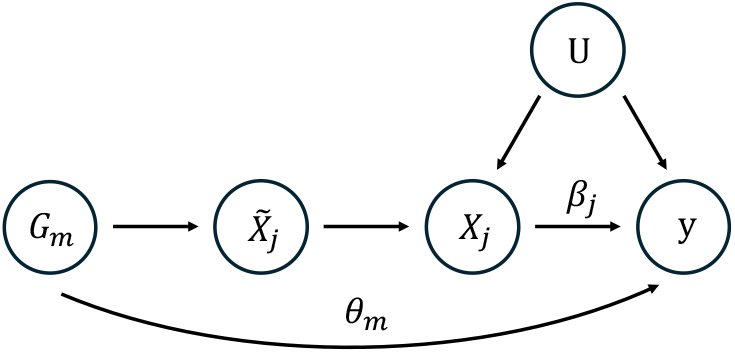
A variant *G*_*m*_ can affect the phenotype *y* through two “path”: one is through molecular trait *X*_*j*_ (effect size *β*_*j*_) and the other is through its direct effect on *y*, i.e. the effect not mediated through *X*_*j*_ (effect size *θ*_*m*_). 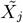 represents the cis-genetic component of molecular trait *X*_*j*_. *U* represents an environmental confounder.

**Figure S2:**
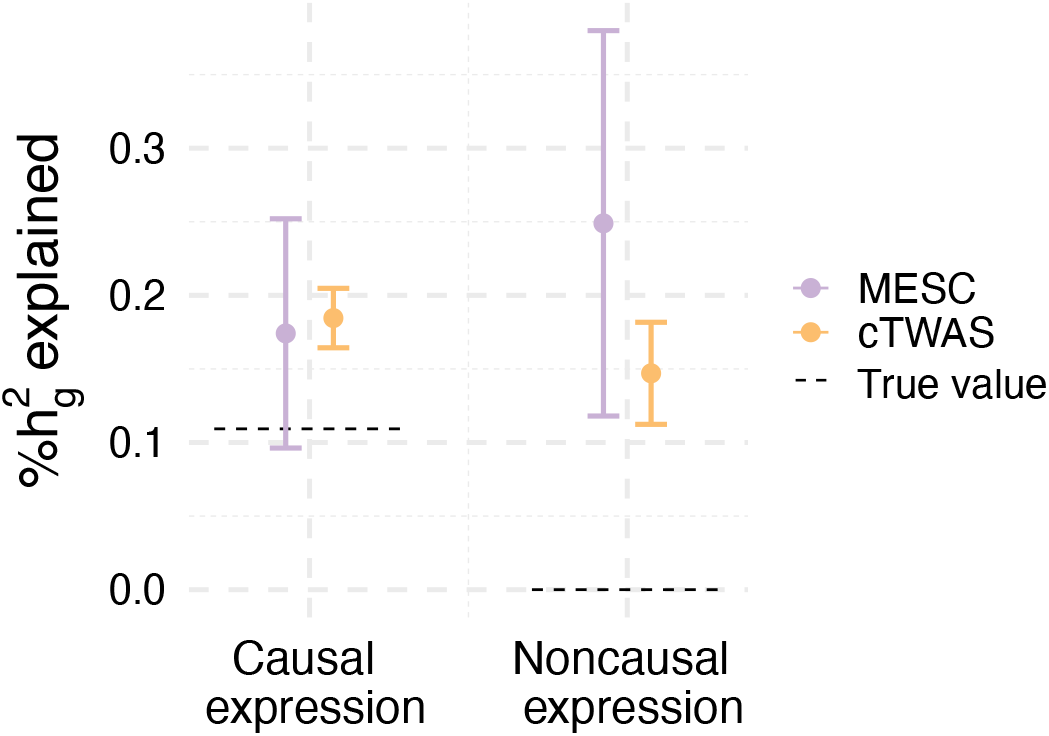
Proportion of heritability explained (PHE) by eQTLs in causal and noncausal tissues estimated by cTWAS and MESC in simulations under the low correlation setting. Dashed horizontal lines indicate the true PHE values. Error bars denote the standard error of the mean.

**Figure S3:**
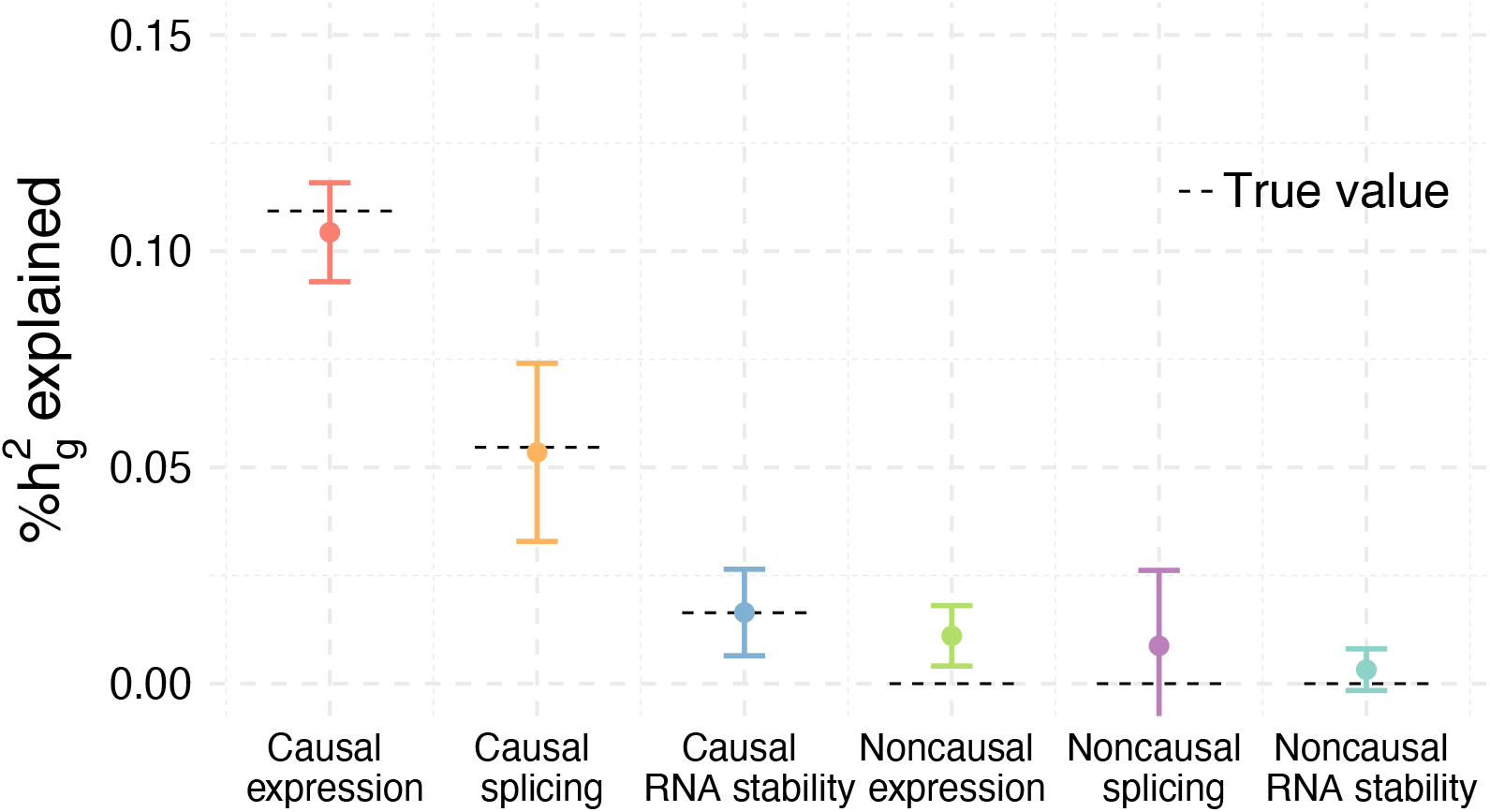
Proportion of heritability explained (PHE) across molecular modalities in causal and noncausal tissues estimated by M-cTWAS under the low correlation setting. Dashed horizontal lines indicate the true PHE values. Error bars denote the standard error of the mean.

**Figure S4:**
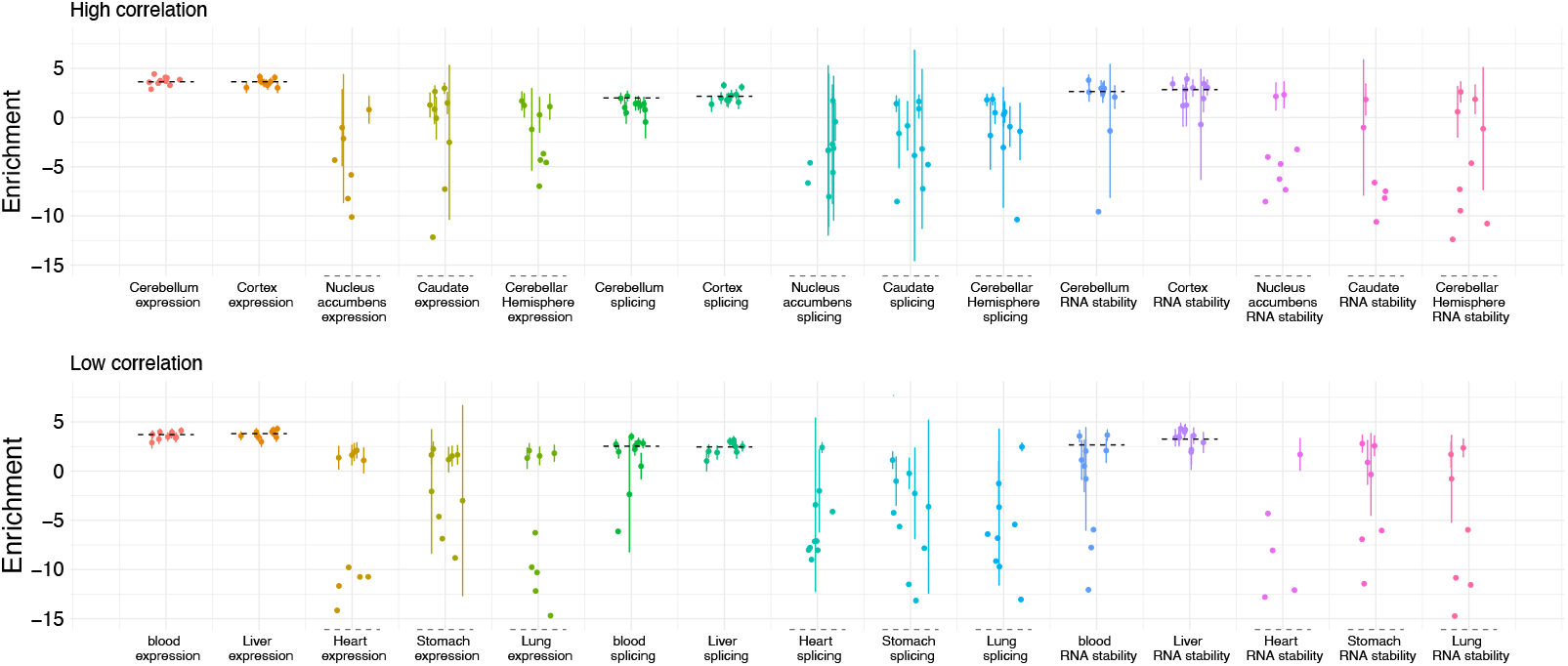
Enrichment of molecular traits estimated by M-cTWAS from simulations (in natural log scale) under the high correlation and low correlation settings. Dashed horizontal lines indicate the true enrichment values. Enrichment is defined as the ratio of prior of molecular traits to prior of SNPs. The error bar shows the standard errors. Note that when there is no signal in molecular traits (at log-scale, enrichment is negative infinity), the estimated enrichment would not be exactly 0, but as long as the enrichment is small (say < 0 at the log-scale), the exact values of the enrichment would not make a difference in practice.

**Figure S5:**
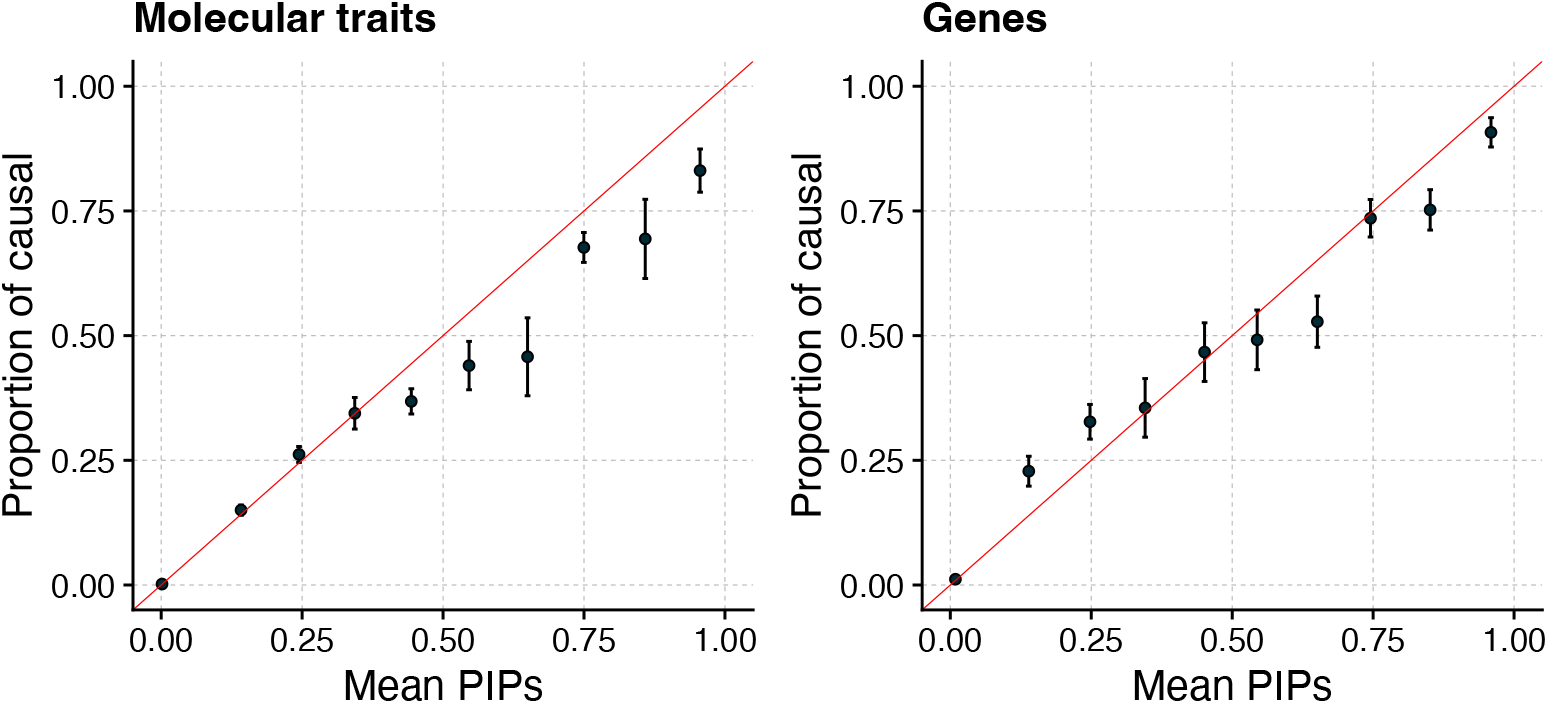
PIP calibration of molecular traits (left) and genes (right) under the low correlation setting. PIPs from all simulations are grouped into bins. The plot shows the proportion of true causal molecular traits or genes (y axis) against the average PIPs (x axis) under each bin. A well-calibrated method should produce points along the diagonal lines (red). Error bars denote the standard error of the mean.

**Figure S6:**
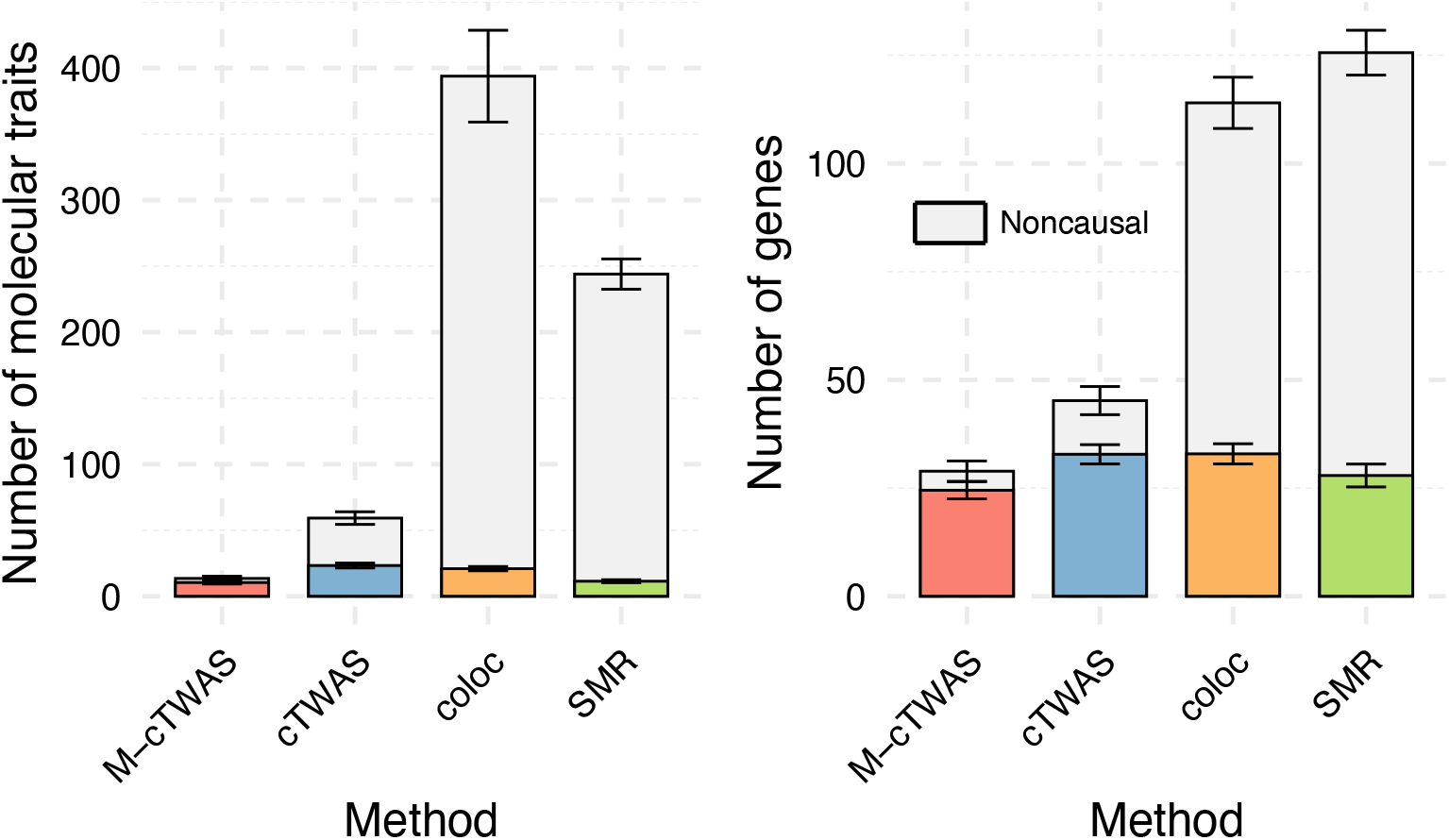
Number of molecular traits (left) and genes (right) identified by M-cTWAS, cTWAS, coloc and SMR under the low correlation setting. We used the following significance thresholds for each method: PIP > 0.8 for M-cTWAS and cTWAS, PP4 > 0.8 for coloc, FDR < 0.05 for SMR with default HEIDI filter. We use different colors for causal molecular traits/genes identified by each method. The top gray bars indicate the noncausal molecular traits/genes. The height of the bar is the mean over ten simulations. Error bars denote the standard error of the mean.

**Figure S7:**
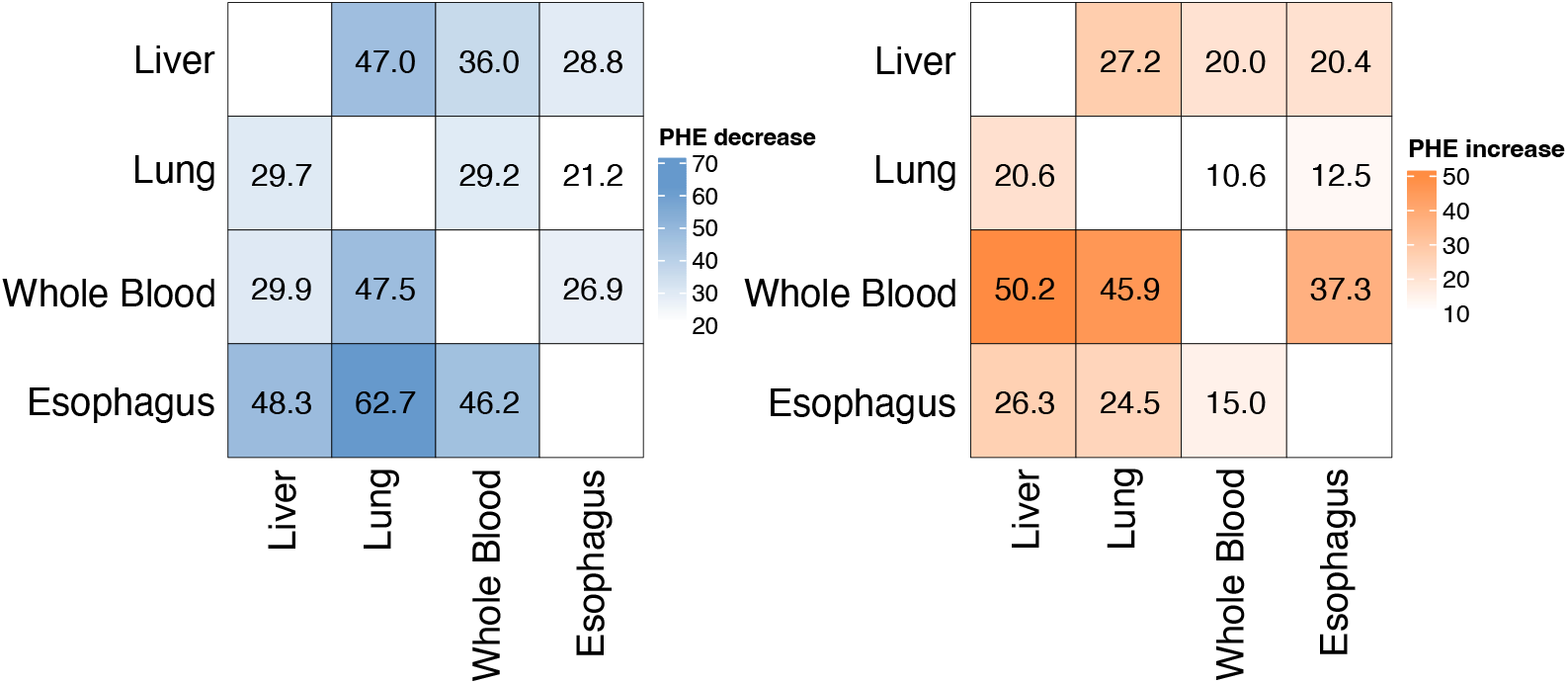
Heatmaps display the percent decrease in PHE attributed to sQTLs in the first tissue after adding sQTLs from a second tissue to the model (left), and the percent increase in total PHE attributed to sQTLs in the joint model compared to the PHE of the first tissue alone (right). Each cell corresponds to an ordered tissue pair, where the row tissue is the “first” tissue and the column tissue is the “second” tissue being added. Results are shown for GWAS of LDL. Esophagus refers to Esophagus Mucosa.

**Figure S8:**
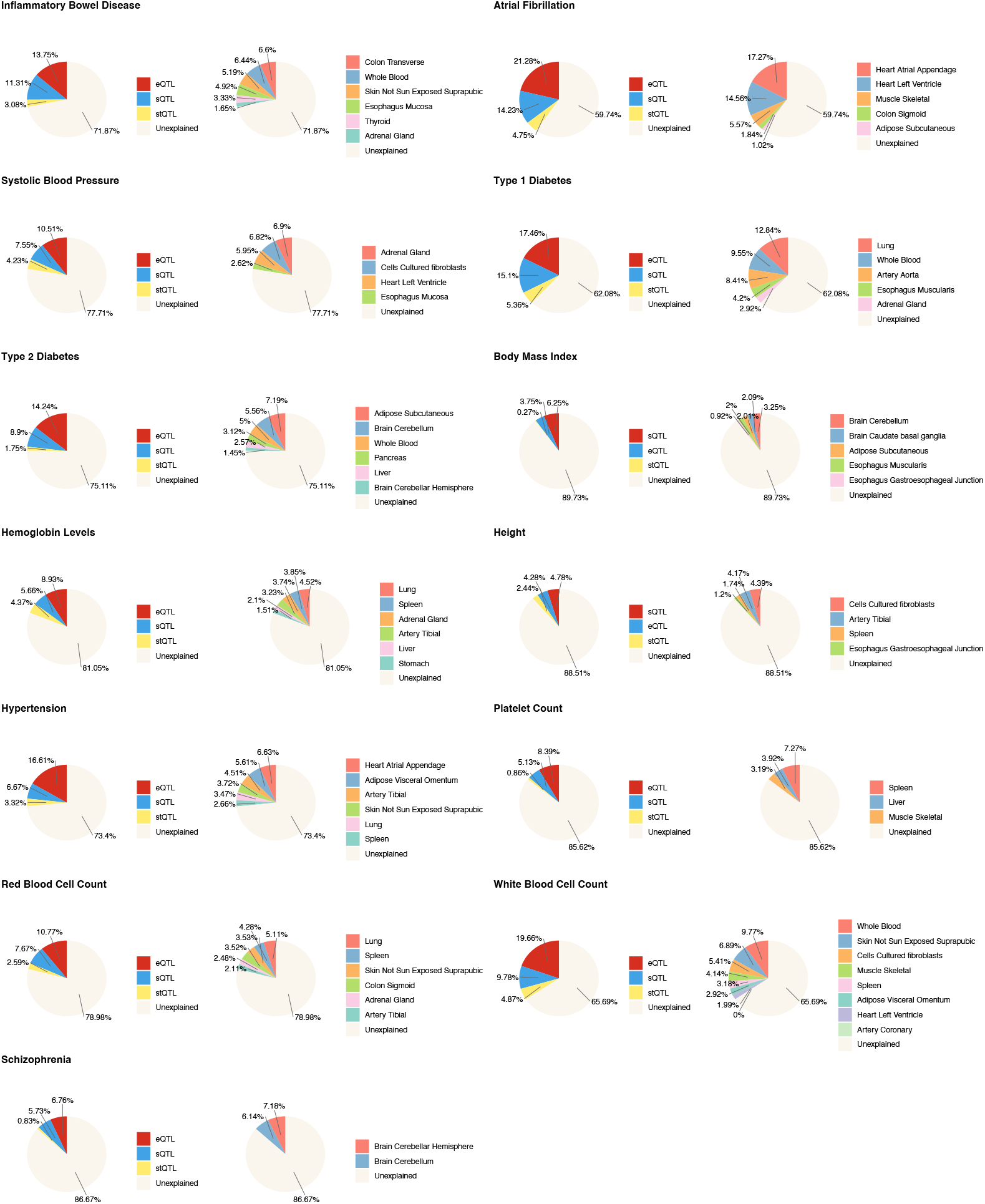
Decomposition of trait heritability by molecular modalities (left) and tissue contexts (right). Results are shown for 13 individual traits.

**Figure S9:**
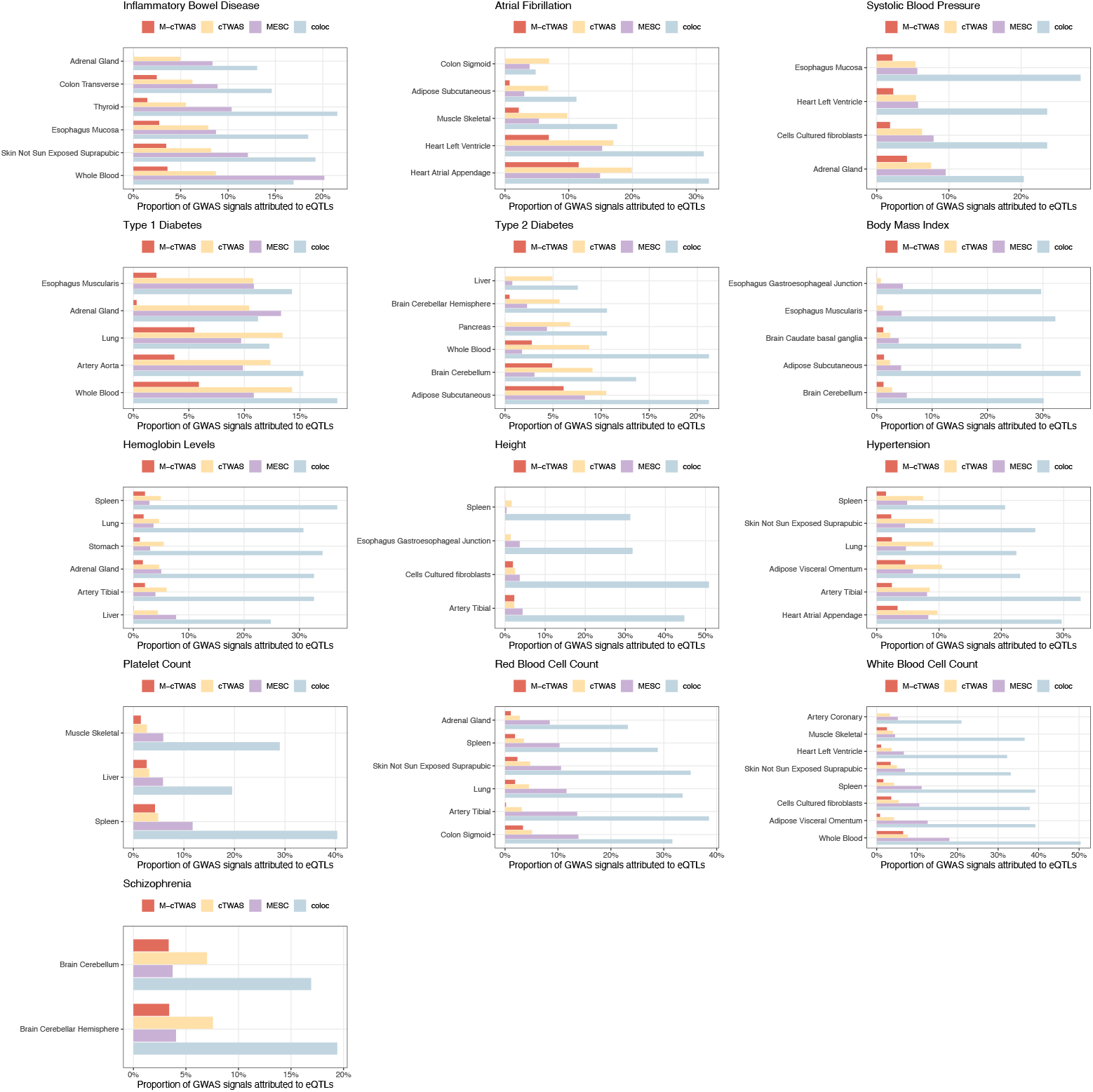
Proportion of heritability explained (PHE) by eQTLs across tissues estimated by M-cTWAS, cTWAS, MESC and coloc. For coloc, we computed the proportion of significant GWAS loci that have colocalized signals with eQTLs. Results are shown for 13 individual traits.

**Figure S10:**
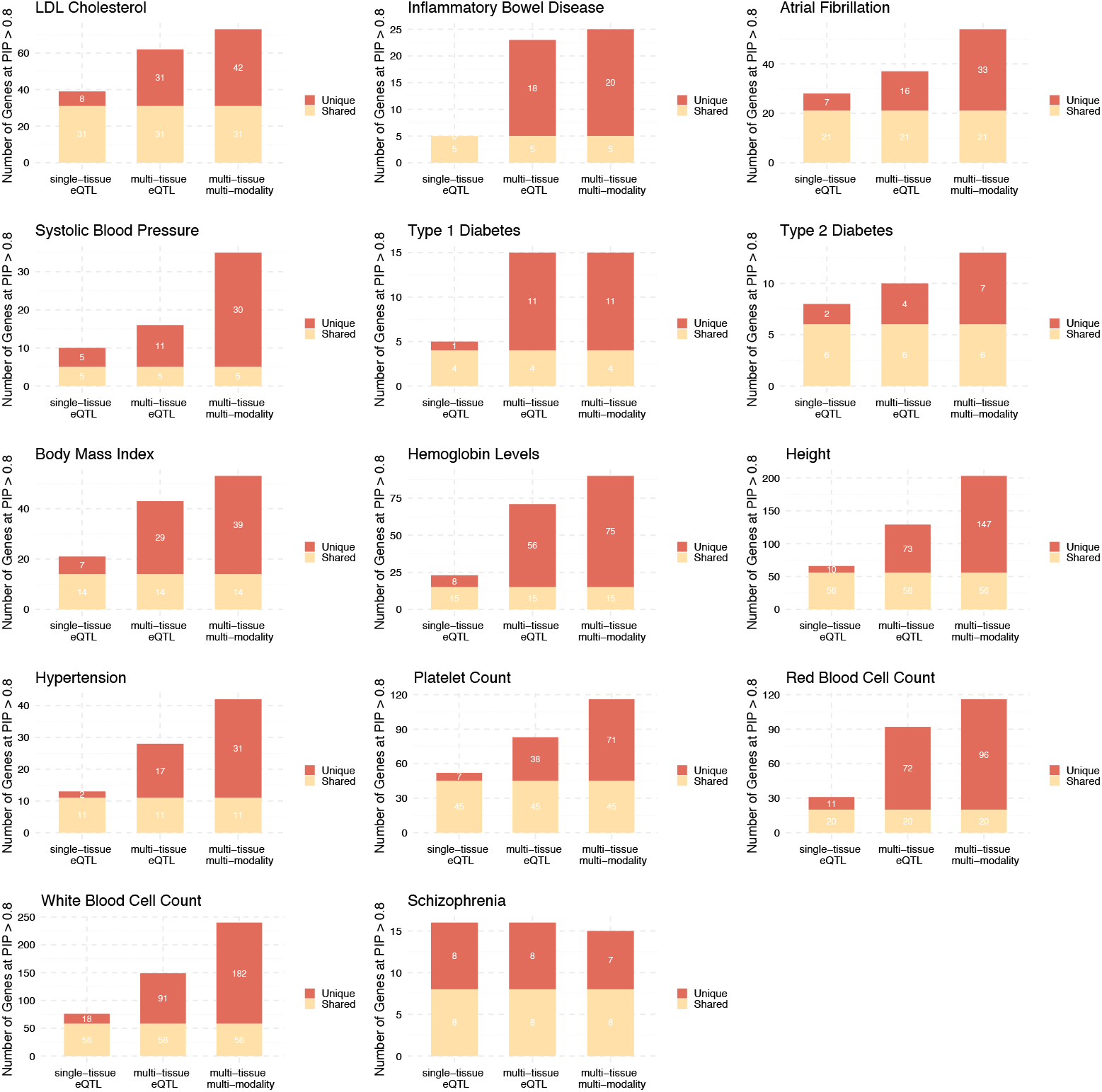
Number of candidate genes identified by M-cTWAS in 14 individual traits from three models: single-tissue eQTL, multi-tissue eQTL and multi-tissue multi-modality. Red color and yellow color indicate unique and shared genes across three models.

**Figure S11:**
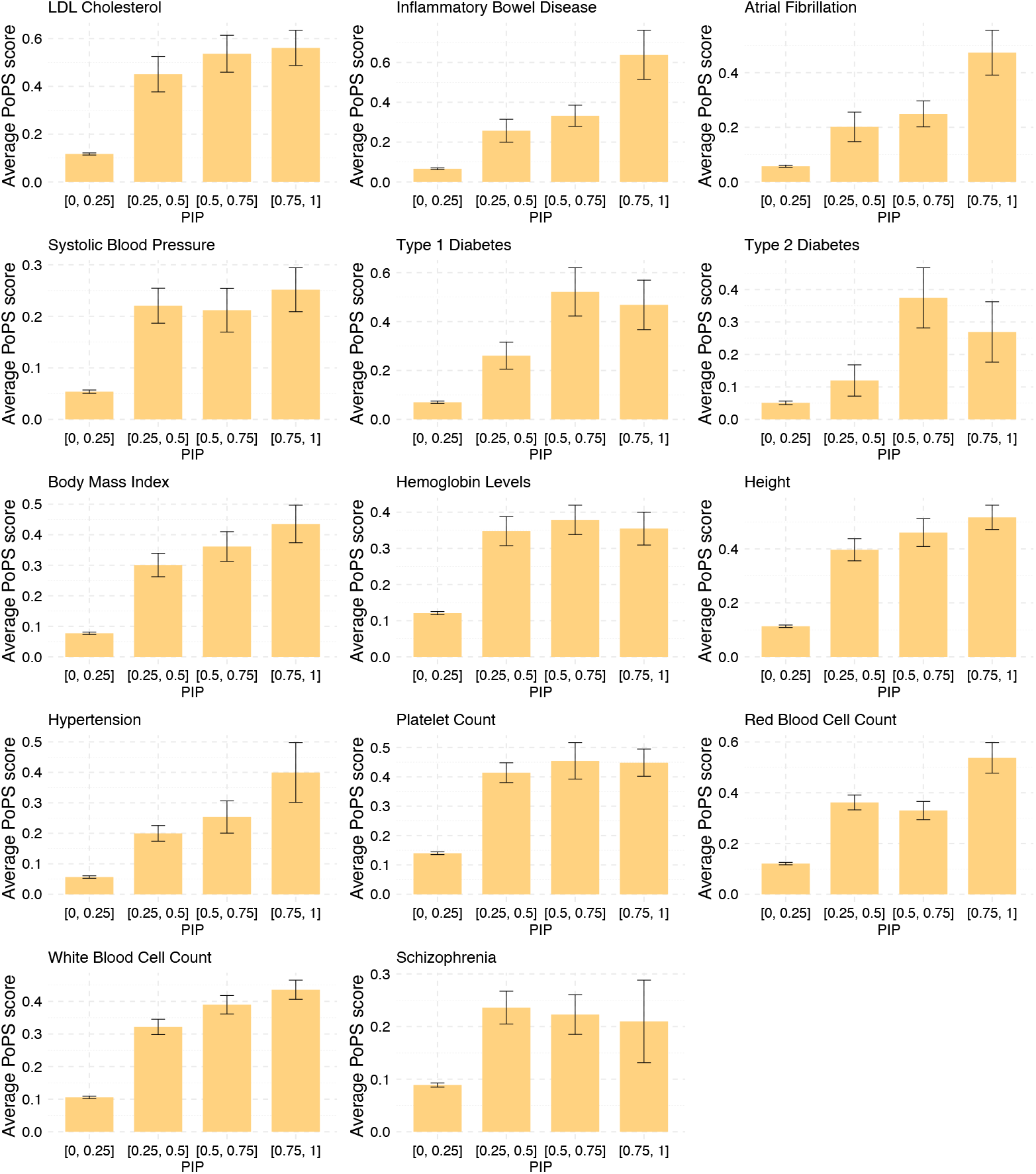
PoPS score (y axis) stratified by M-cTWAS posterior inclusion probability (PIP) bins in 14 individual traits. Bars show the average PoPS across genes within each bin. Error bars denote the standard error of the mean.

**Figure S12:**
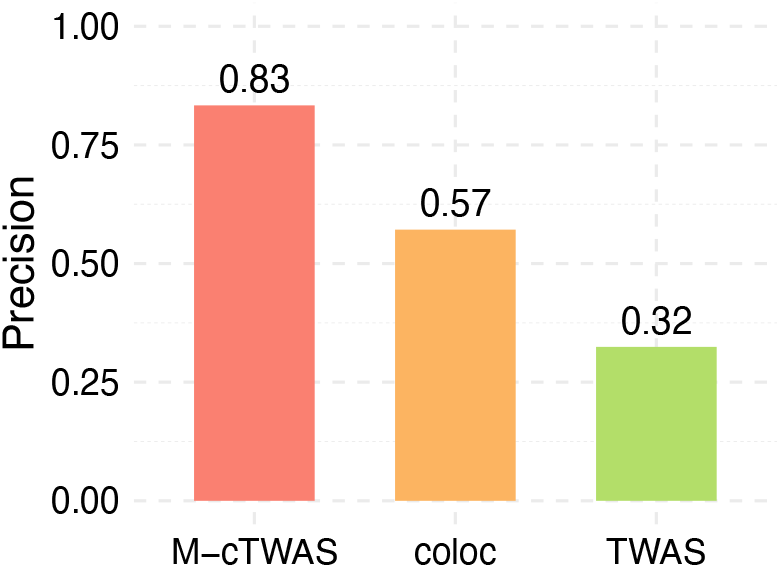
Precision of M-cTWAS, coloc and TWAS in distinguishing IBD high-confidence risk genes from nearby bystander genes in the same loci.

**Figure S13:**
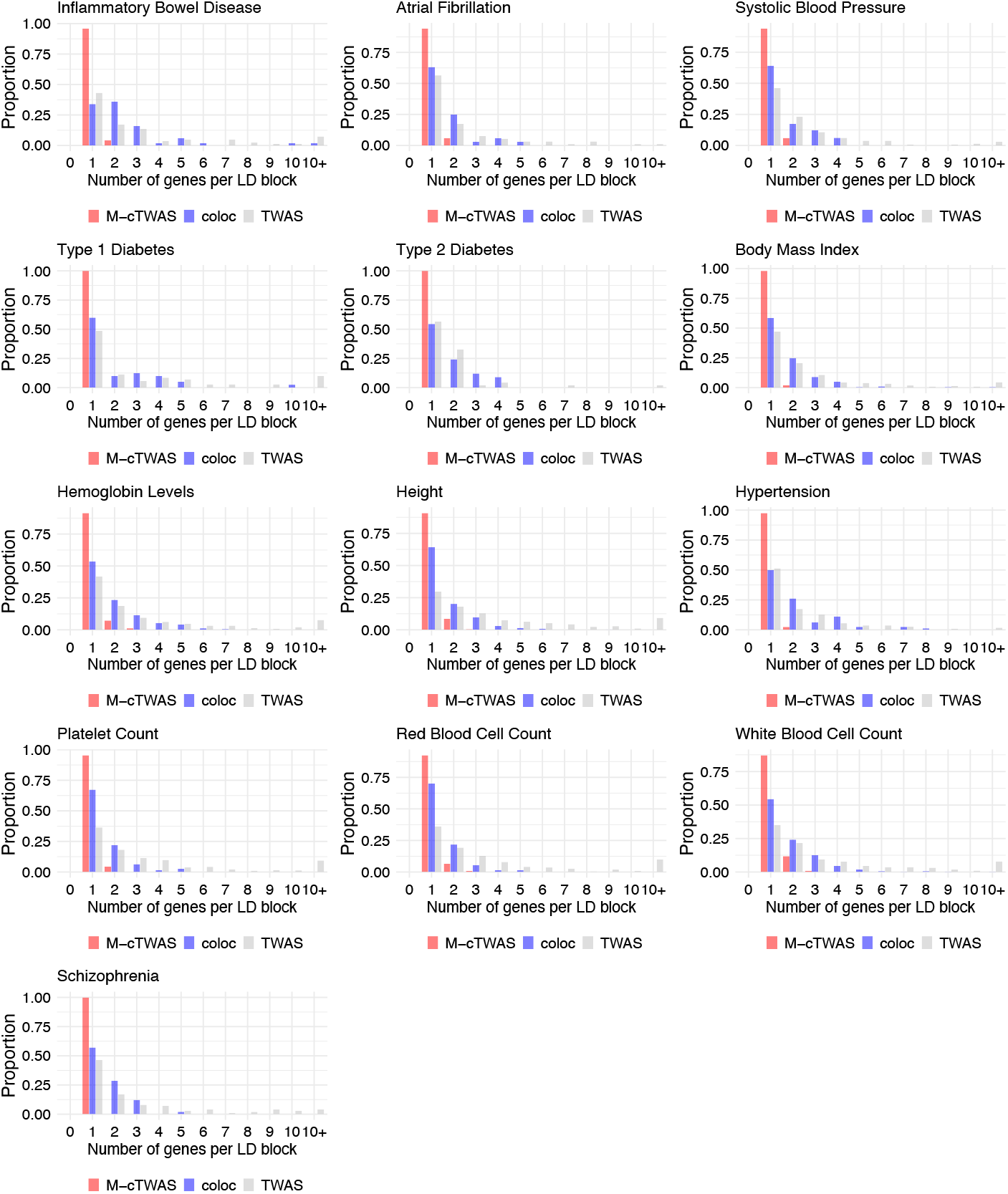
The distribution of the number of genes identified by M-cTWAS, coloc and TWAS per LD block in GWAS of 13 individual traits.

**Figure S14:**
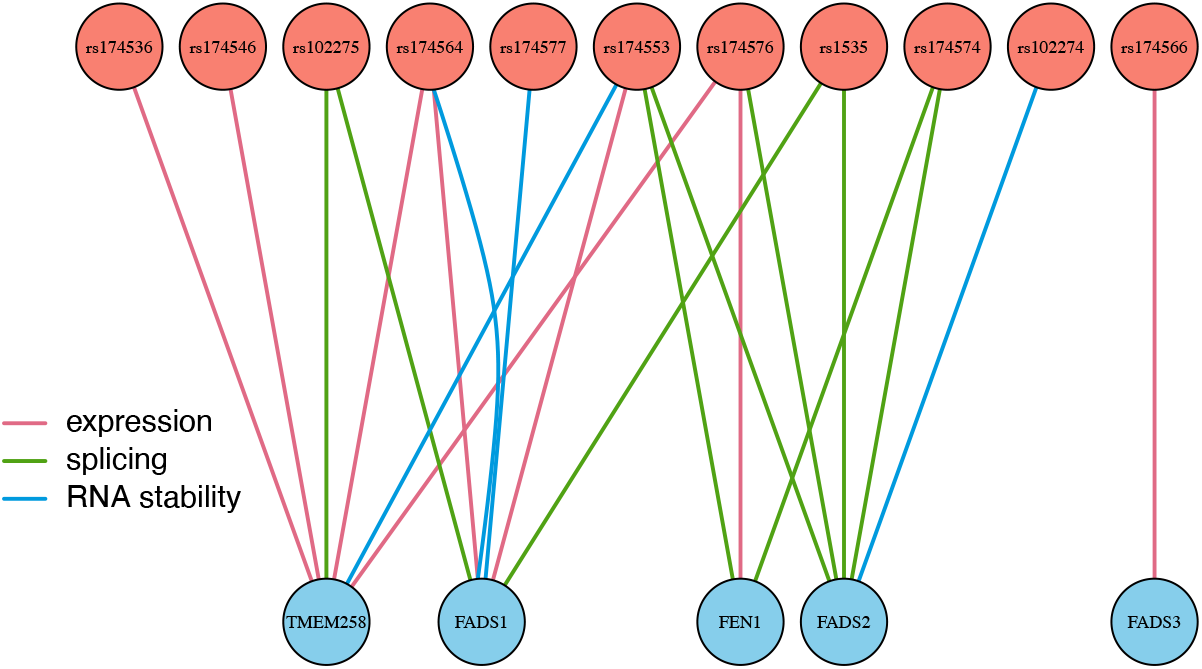
Network of genes (lower circles) prioritized by coloc (PP4 > 0.8) and their supporting SNPs (upper circles) at the FADS1 locus of LDL. Edges indicate molecular mechanisms that supporting SNPs act through. Colors of edges indicate different types of molecular mechanisms.

**Figure S15:**
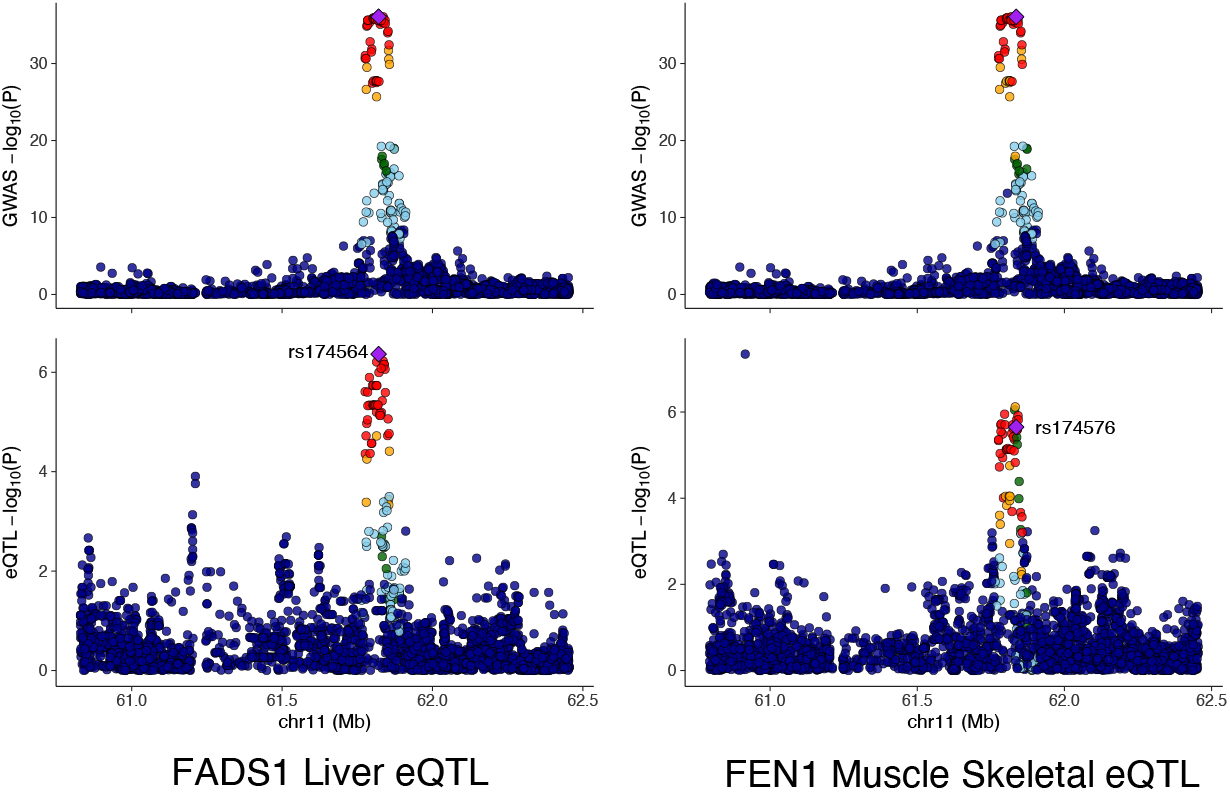
Colocalization of eQTL and GWAS signals. The colocalized SNP is marked on the plot and colored in purple. Color indicates the LD of each SNP with the colocalized SNP. Shown are two out of the five colocalized examples (as listed in Figure S14): Liver eQTLs of FADS1 and muscle skeletal eQTLs of FEN1 are colocalized with the GWAS signal in this locus: Liver eQTLs of *FADS1* and muscle skeletal eQTLs of *FEN1* are colocalized with the GWAS signal in this locus.

**Figure S16:**
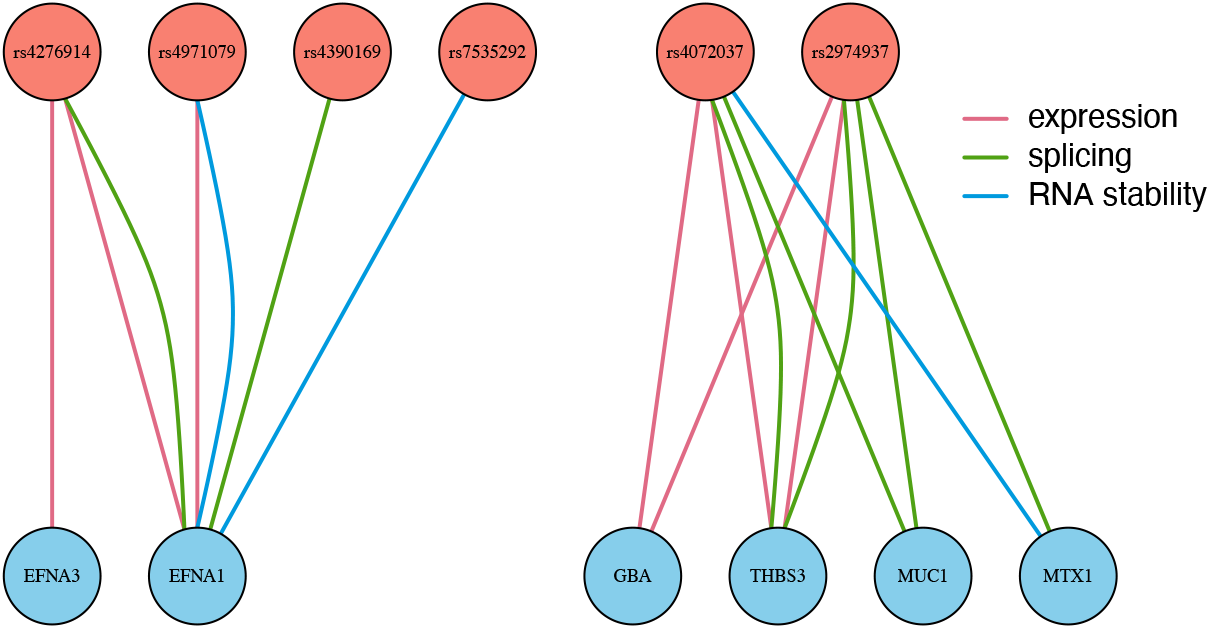
Network of genes (lower circles) prioritized by coloc (PP4 > 0.8) and their supporting SNPs (upper circles) at the ADAM15 locus in IBD. Edges indicate molecular mechanisms that supporting SNPs act through. Colors of edges indicate different molecular mechanisms. ADAM15 itself has coloc PP4 < 0.8.

**Figure S17:**
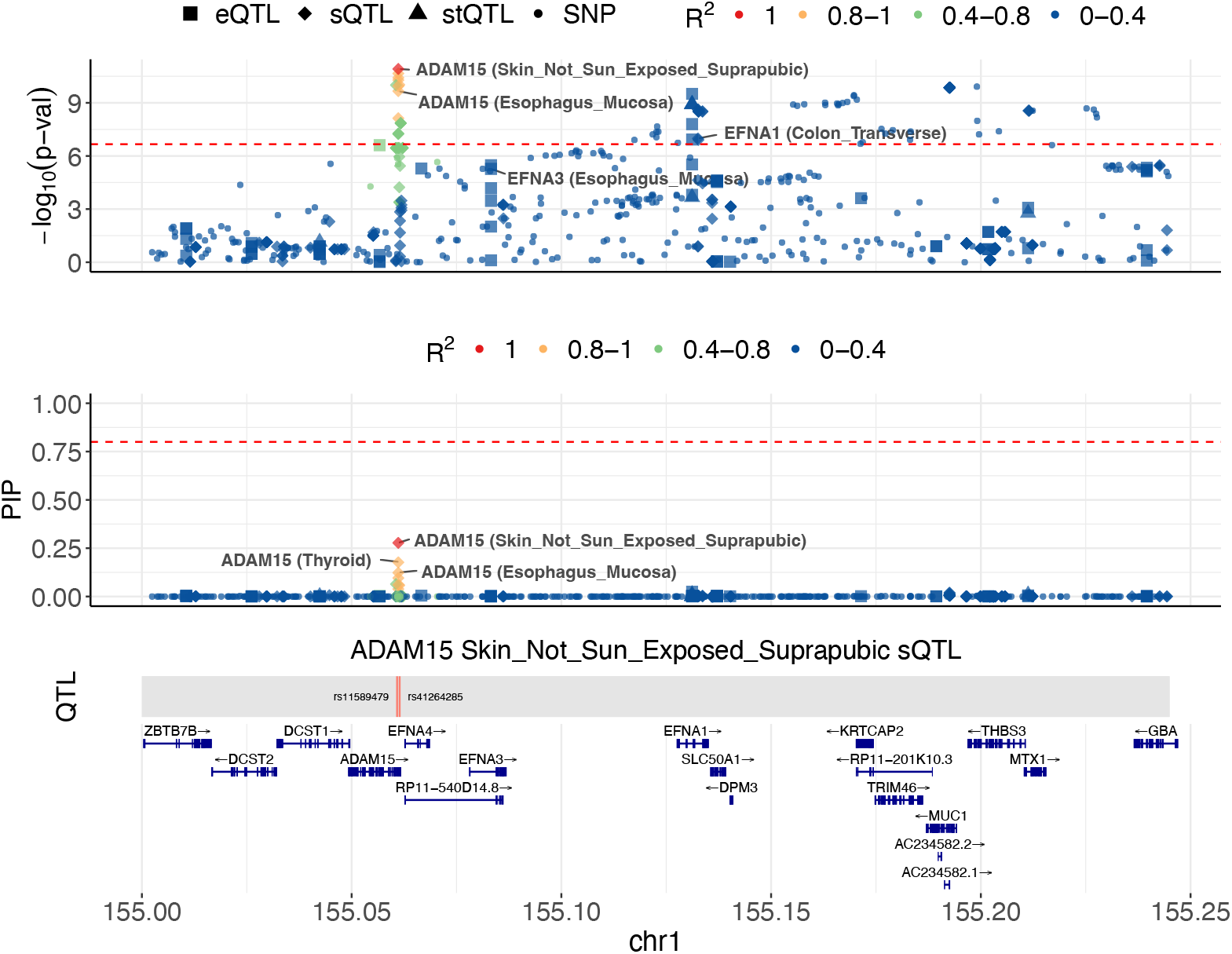
Locus plot of the *ADAM15* gene locus. The upper panel shows associations of molecular traits and variants with the trait (LDL). The middle panel shows M-cTWAS PIPs of molecular traits and variants. The combined PIP of ADAM15 is 0.915. The lower two panels show the genomic location of the QTLs (weights in prediction model) of the focal molecular trait and the gene track. Different molecular modalities are shown in different shapes. Colors indicate the LD with the focal molecular trait.

**Figure S18:**
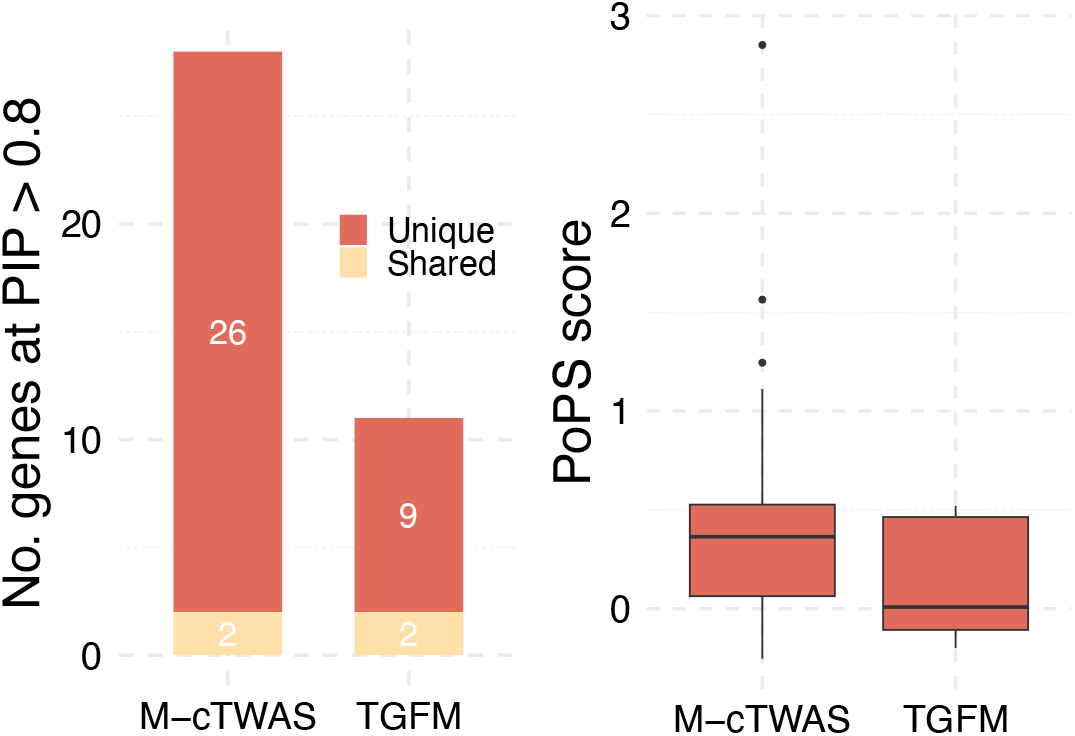
Number of genes identified by M-cTWAS and TGFM in GWAS of hypertension (HTN) at the PIP > 0.8 threshold (left). Colors indicate unique and shared genes identified by two methods. PoPS scores of genes uniquely identified by M-cTWAS and TGFM (right).

**Figure S19:**
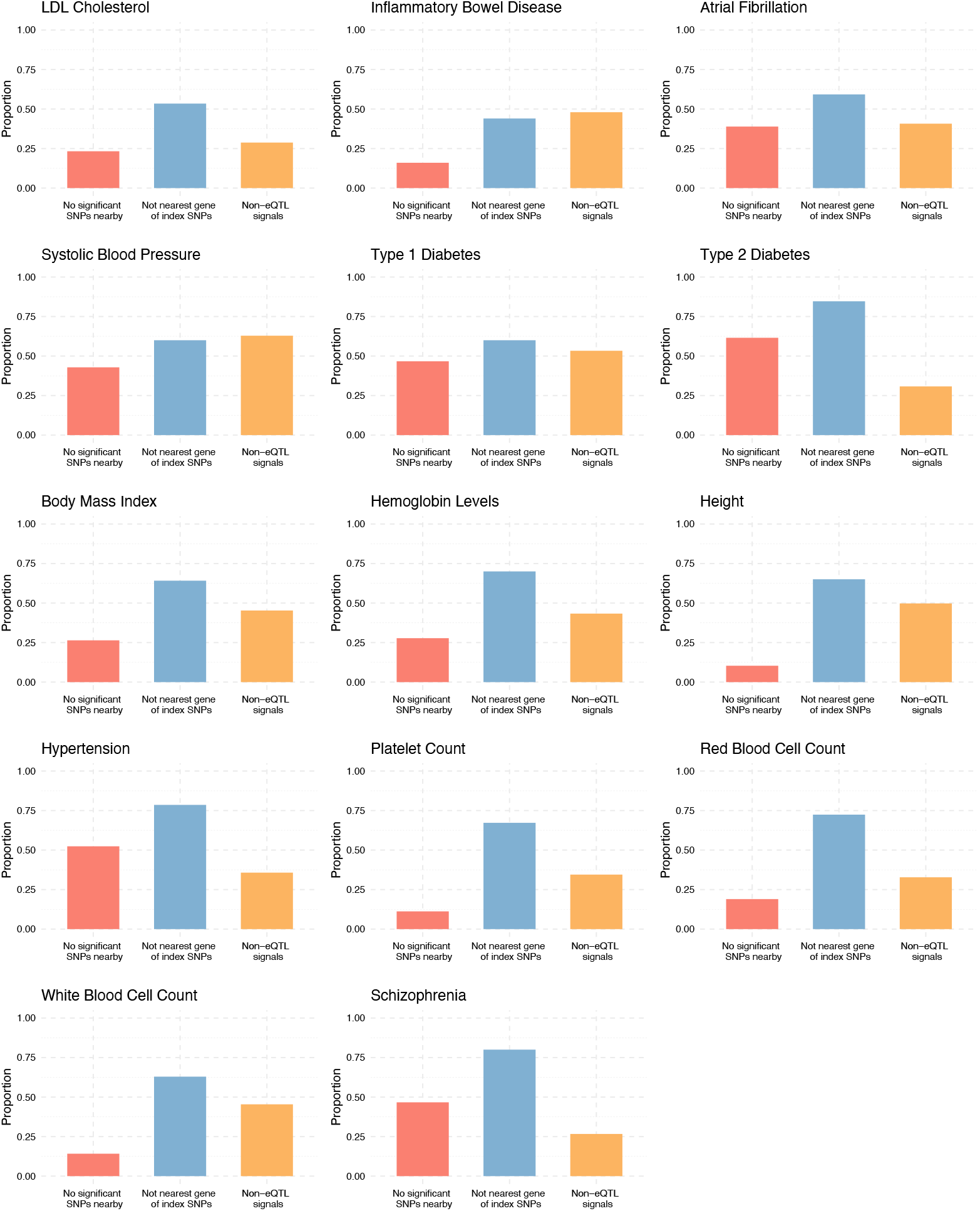
The proportion of genes identified by M-cTWAS in 14 individual traits that fall into three novel gene categories (i) no significant SNPs nearby: no genome-wide significant SNP (*P* < 5 × 10^−8^) within *±*500 kb of the gene’s TSS; (ii) not nearest gene of GWAS loci: not the nearest gene to the GWAS lead SNP in a locus; and (iii) non-eQTL signals: genes detected exlusively through splicing and RNA stability.

**Figure S20:**
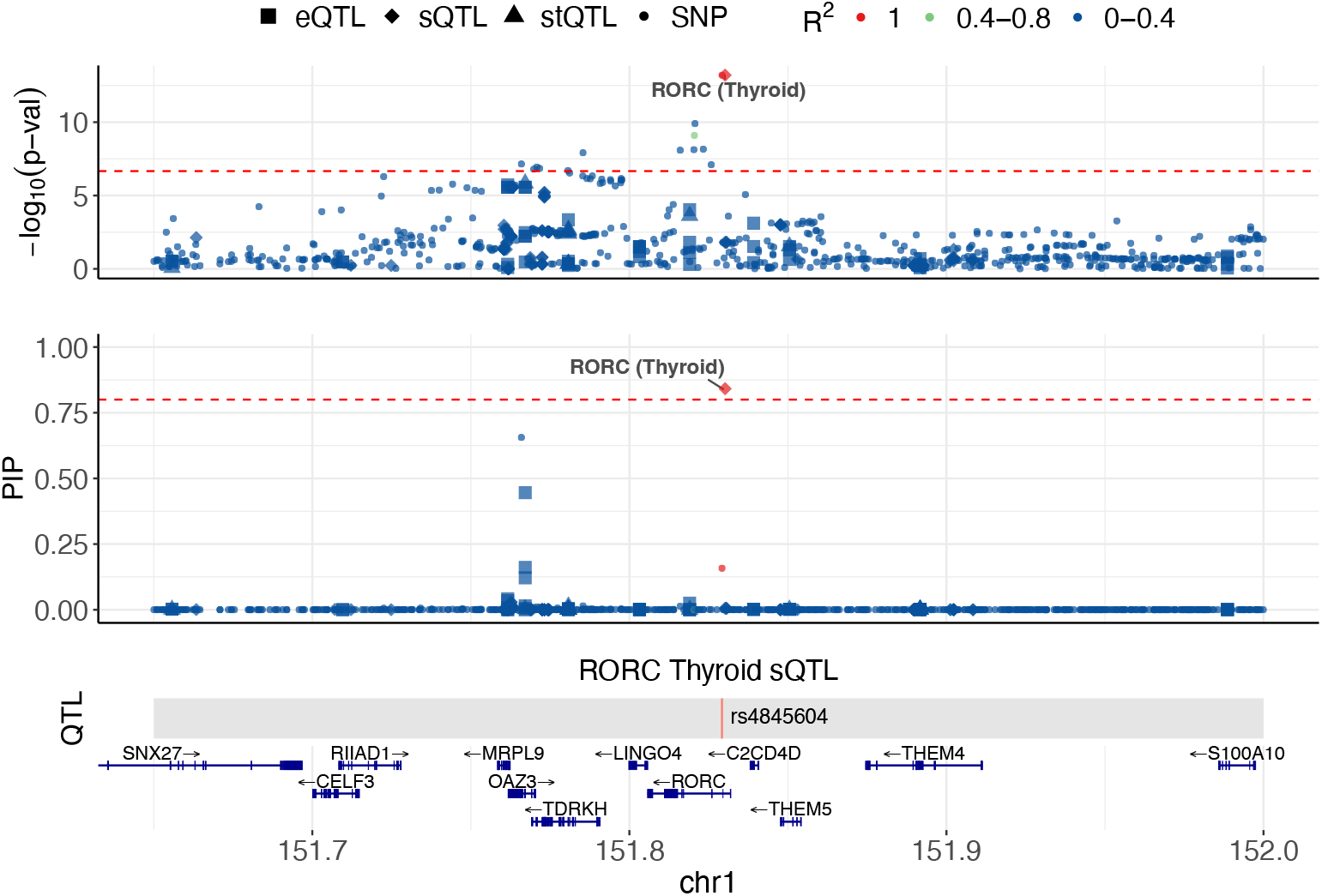
Locus plot of the *RORC* gene locus. In each locus, the upper panel shows associations of molecular traits and variants with the trait (LDL). The middle panel shows M-cTWAS PIPs of molecular traits and variants. The lower two panels show the genomic location of the QTLs (weights in prediction model) of the focal molecular trait and the gene track. Different molecular modalities are shown in different shapes. Colors indicate the LD with the focal molecular trait.

**Figure S21:**
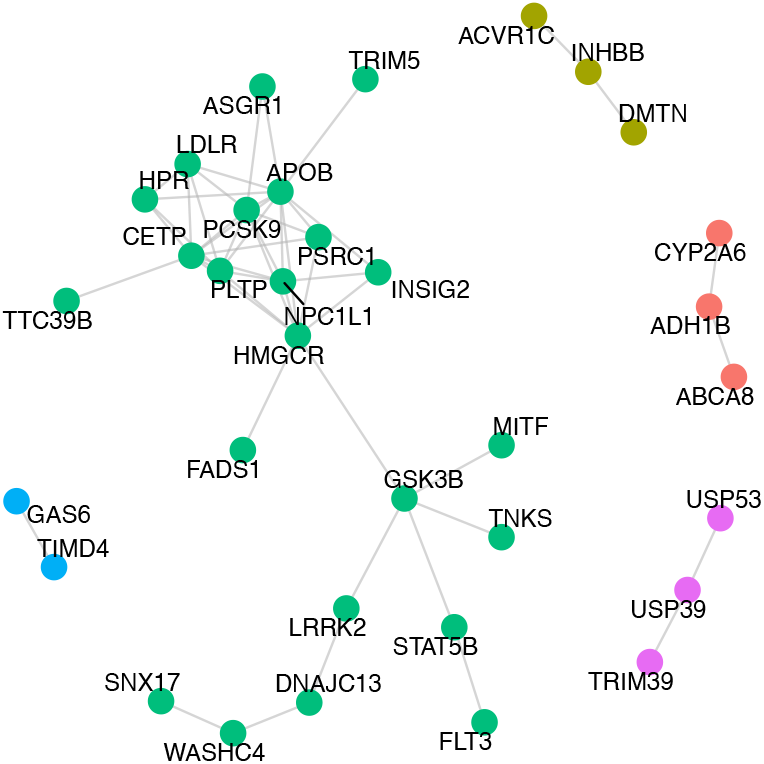
String interaction network of LDL candidate genes identified by M-cTWAS (PIP > 0.8). Colors indicate cluster memberships from *k*-means clustering (*k*=5).

**Figure S22:**
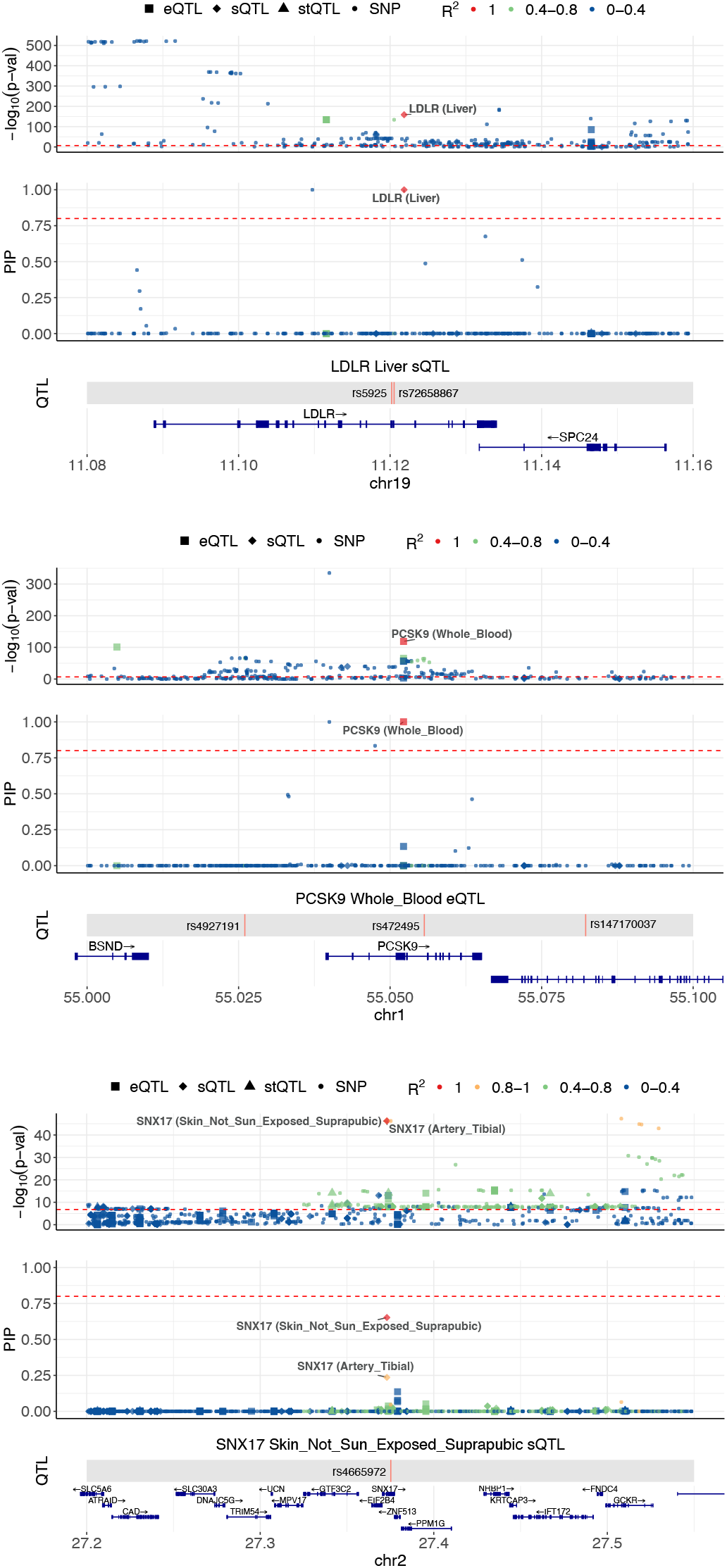
Locus plot of the *LDLR* (upper), *PCSK9* (middle) and *SNX17* (lower) gene locus. The upper panel shows associations of molecular traits and variants with the trait (LDL). The middle panel shows M-cTWAS PIPs of molecular traits and variants. The lower two panels show the genomic location of the QTLs (weights in prediction model) of the focal molecular trait and the gene track. Different molecular modalities are shown in different shapes. Colors indicate the LD with the focal molecular trait.

**Figure S23:**
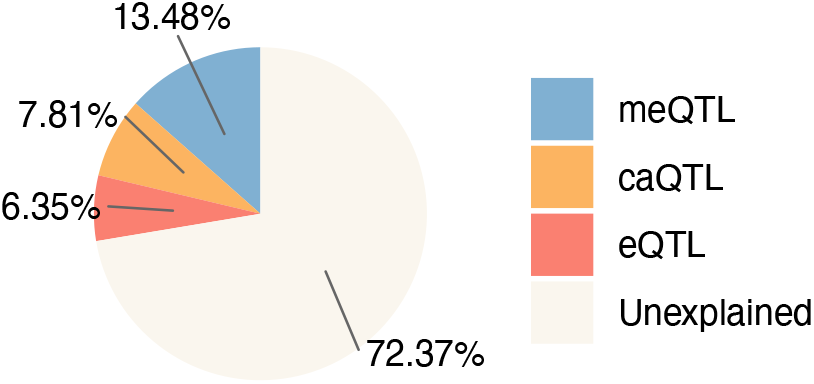
Partition of schizophrenia (SCZ) heritability across brain DNA methylation, brain chromatin accessibility and brain gene expression (PsychENCODE).

**Figure S24:**
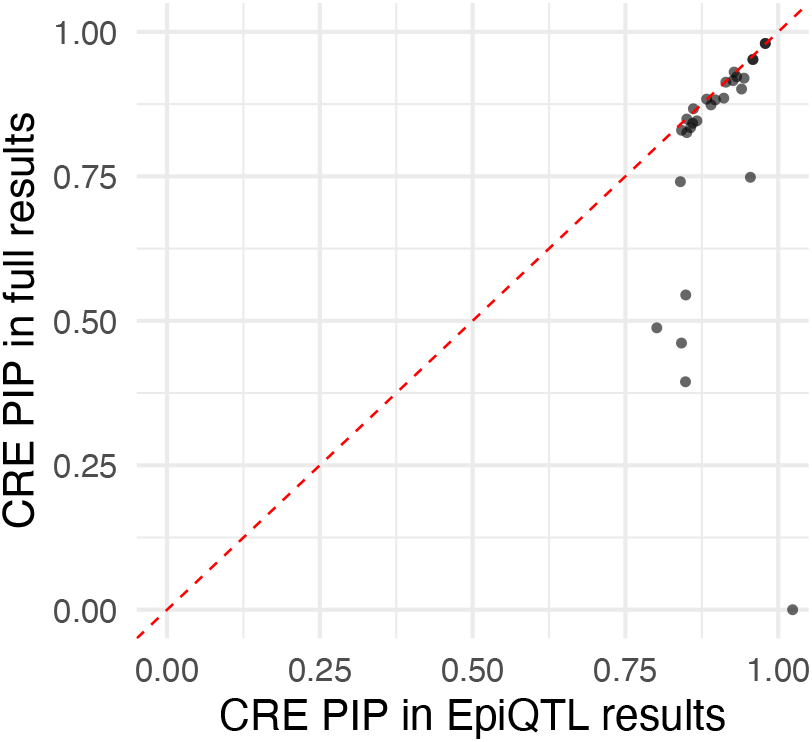
PIPs of candidate CREs in the M-cTWAS analysis with epiQTLs alone (X-axis) compared to those in the jointly M-cTWAS analysis with epiQTLs and eQTLs (Y-axis).

**Figure S25:**
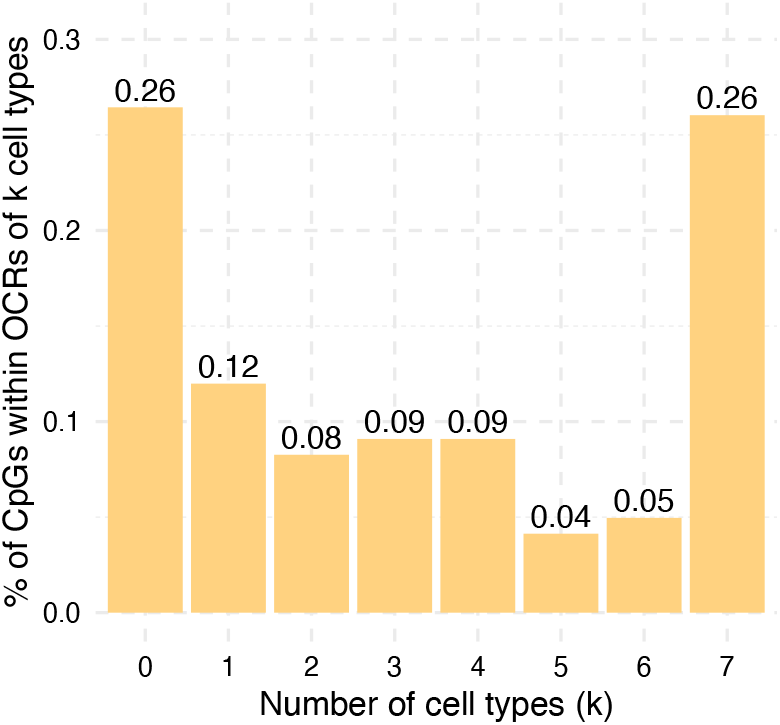
Distribution of the proportion of CpGs located within open chromatin regions (OCRs) of *k* cell types.

**Figure S26:**
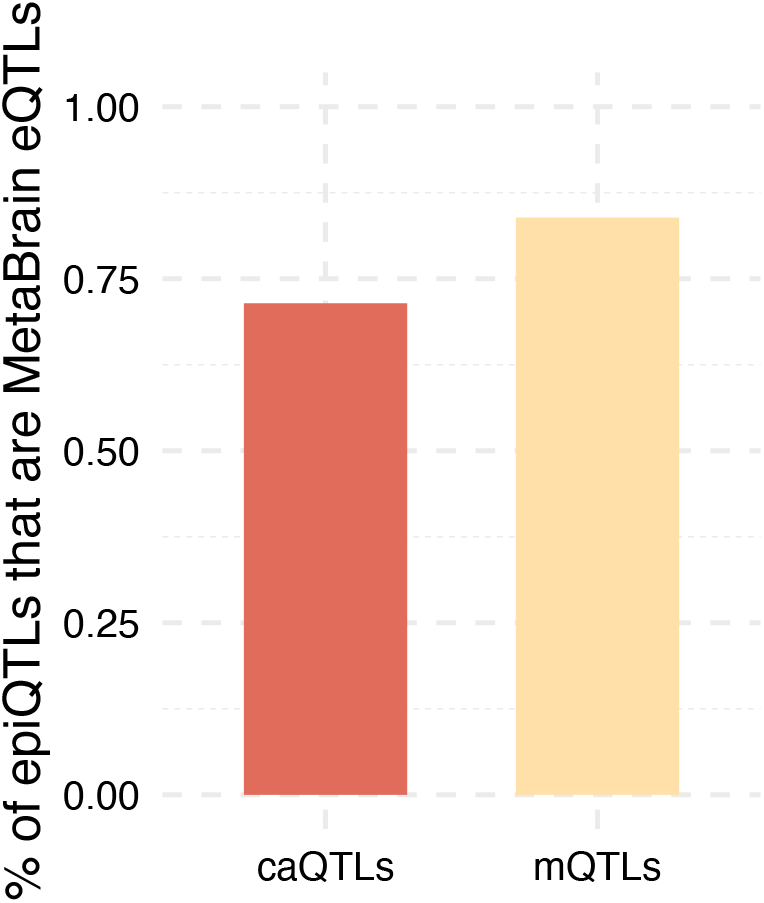
The proportion of caQTLs and meQTLs that are identified as eQTLs in the MetaBrain study.

